# Global Economic Cost of Deaths Attributable to Ambient Air Pollution: Disproportionate Burden on the Ageing Population

**DOI:** 10.1101/2020.04.28.20083576

**Authors:** Hao Yin, Michael Brauer, Junfeng (Jim) Zhang, Wenjia Cai, Ståle Navrud, Richard Burnett, Courtney Howard, Zhu Deng, Daniel M. Kammen, Hans Joachim Schellnhuber, Kai Chen, Haidong Kan, Zhanming Chen, Bin Chen, Ning Zhang, Zhifu Mi, D’Maris Coffman, Yiming Wei, Aaron Cohen, Dabo Guan, Qiang Zhang, Peng Gong, Zhu Liu

## Abstract

**Background:** The health impacts of ambient air pollution impose large costs on society. While all people are exposed to air pollution, older individuals tend to be disproportionally affected. As a result, there is growing concern about the public health impacts of air pollution as many countries undergo rapid population ageing. We investigated the spatial and temporal variation in the health economic cost of deaths attributable to ambient air pollution, and its interaction with population ageing from 2000 to 2016 at global and regional levels.

**Methods:** We developed an age-adjusted measure of the value of a statistical life year (VSLY) to estimate the health economic cost attributable to ambient PM_2.5_ pollution using the Global Burden of Disease 2017 data and country-level socioeconomic information. First, we estimated the global age- and cause-specific mortality and years of life lost (YLL) attributable to PM_2.5_ pollution using the global exposure mortality model (GEMM) and global estimates of exposure derived from ground monitoring, satellite retrievals and chemical transport model simulations at 0.1° × 0.1° (~11 km at the equator) resolution. Second, for each year between 2000 and 2016, we translated the YLL within each age-group into a health-related economic cost using a country-specific, age-adjusted measure of VSLY. Third, we decomposed the major driving factors that contributed to the temporal change in health costs related to PM_2.5_. Finally, we conducted a sensitivity test to analyze the variability of the estimated health costs to four alternative valuation measures. We identified the uncertainty intervals (UIs) from 1000 draws of the parameters and exposure-response functions by age, cause, country and year. All economic values are reported in 2011 purchasing-power-parity-adjusted US dollars.

**Findings:** Globally, 8.42 million (95% UI: 6.50, 10.52) deaths and 163.68 million (116.03, 219.44) YLL were attributable to ambient PM_2.5_ in 2016. The average attributable mortality for the older population was 12 times higher than for those younger than 60 years old. In 2016, the global health economic cost of ambient PM_2.5_ pollution for the older population was US$2.40 trillion (1.89, 2.93) accounting for 59% of the cost for the total population. The health cost for the older population alone was equivalent to 2.1% (1.7%, 2.6%) of global gross domestic product (GDP) in 2016. While the economic cost per capita for the older population was US$2739 (2160, 3345) in 2016, the cost per capita for the younger population was only US$268 (205, 335). From 2000 to 2016, the annual global health economic cost for the total population increased from US$2.37 trillion (1.88, 2.87) to US$4.09 trillion (3.19, 5.05). Decomposing the factors that contributed to the rise in health economic costs, we found that increases in GDP per capita, population ageing, population growth, age-specific mortality reduction, and PM_2.5_ exposure changed the total health economic cost by 77%, 21.2%, 15.6%, -41.1% and -0.2%, respectively. Compared to using an age-invariant VSLY or an age-invariant value of a statistical life (VSL), the estimates of the older population’s share of the total health economic cost using an age-adjusted VSLY was 2 and 18 percentage points lower, respectively.

## Interpretation

The health economic cost borne by the older population almost doubled between 2000 and 2016, driven primarily by GDP growth, population ageing and population growth. Compared to younger individuals, air pollution leads to disproportionately higher health costs amongst the older population, even after accounting for their relatively shorter remaining life expectancy and increased disability. The age-specific estimates of health economic cost inform the optimal design of air pollution reduction strategies and allocation of healthcare resources. The positive relationship between age and economic costs suggests that countries with severe air pollution and rapid aging rates would particularly benefit from improving their air quality. In addition, strategies aimed at enhancing healthcare services, especially for the older population, may be beneficial for reducing the costs of ambient air pollution.

## I Background

As a leading health risk factor, ambient air pollution imposes enormous costs on society.^2,3^ Worldwide exposure to ambient PM_2.5_ results in 4.2 million to 8.9 million deaths.^4,5^ By reducing life expectancy, air pollution causes substantial loss of human capital, productivity, and social wellbeing. The negative health effects of air pollution are especially harmful to older people.^6^ According to the United Nations, the population aged 60 years and older has grown globally by 50% from 2000 to 2016.^7^ Not only has the shift in age demographics meant that the population is now more vulnerable to air pollution, but in many countries, the ageing population is concentrated in regions with rising air pollution.^3^ Thus, an expanding elderly population can potentially amplify the health costs of exposure to air pollution on social welfare, jeopardizing the health targets outlined by the United Nations’ Sustainable Development Goals.

To inform policy decisions regarding the mitigation of air pollution, a growing literature has evaluated the economic costs of the effect of air pollution on human health. These studies have documented large health costs associated with air pollution on both global and regional scales.^8–10^ According to a World Bank report on the cost of air pollution, the health economic cost of ambient PM_2.5_ was US$3.55 trillion in 2013, equivalent to 3.5% of global GDP.^8^ Recently, the 2019 *Lancet* Countdown on Health and Climate Change added the health costs of air pollution as an indicator of the health co-benefits of climate change mitigation.^11^ The burning of fossil fuels was responsible for much of the burden of air pollution, which was associated with 1.1 years of lost life expectancy.^12^ As activities related to fuel combustion also produce greenhouse gases, valuations of the costs of air pollution provide compelling rationale for decision-makers to make changes that not only reduce air pollution, but also the drivers of climate change.

Although the negative health effects of air pollution increase with age, previous studies adopted measures of the value of a statistical life (VSL) or value of a statistical life year (VSLY) that were constant across all age groups to quantify the cost of air pollution attributable deaths. For instance, the *Lancet* Commission on Investing in Health outlined a methodology to value the changes in mortality by assuming that VSL was proportional to remaining life expectancy.^13^ Estimates derived from this methodology do not account for the theoretical and empirical evidence that both VSL and VSLY vary with people’s age due to changes in remaining life expectancy, life quality and socioeconomic status.^14–16^ Therefore, applying an age-invariant VSL or VSLY inaccurately estimates the health economic cost of air pollution, which might bias the optimal allocation of resources for pollution control and public health investments.

In this study, we applied an age-adjusted VSLY to investigate the contribution of population ageing to the global health economic cost of deaths attributable to ambient PM_2.5_ pollution. By analyzing the interaction between ambient air pollution and population ageing, the findings from this study will inform the design of pollution control policies and healthcare investment strategies to improve the public health response to population ageing.

## Research in context

### Evidence before this study

We searched Web of Science, Google Scholar, and publicly available literature up to March 2020 for the terms “air pollution”, “mortality”, “health cost” and “population ageing” without language restrictions to find studies that examine the relationship between population ageing and the health economic cost of air pollution. Previous research found that population ageing was a major driver of the substantial growth in global non-communicable diseases. However, although many studies have assessed the health costs of air pollution, few have considered how it changed as a result of population aging. We found multiple articles, including reports from the World Bank, the World Health Organization (WHO) and the Organization for Economic Co-operation and Development (OECD) that estimated the health cost of air pollution at both regional and global scales. These studies applied either an age-invariant value of a statistical life (VSL) or an age-invariant value of a statistical life year (VSLY) as their economic measure of attributable mortality. For example, using an age-invariant VSL measure, the World Bank calculated that the total welfare loss from deaths attributable to ambient air pollution rose from US$2.18 trillion to US$3.55 trillion between 1990 and 2013. By assuming an age-invariant VSL or VSLY, these studies do not account for the elevated risks and fewer life years lost that older people face when exposed to air pollution and are unable to determine the effect of population ageing on its economic cost.

### Added value of this study

This study examines the interaction between population ageing and the global health cost of deaths attributable to ambient air pollution. Unlike previous studies that adopted an age-invariant VSL or VSLY to estimate the health cost of air pollution, we developed an age-adjusted measure of VSLY that incorporates the effects of variations in life expectancy, wealth distribution and life quality over the lifecycle. By accounting for these factors, our methodology provides an improved measure for the value of mortality abatement. Additionally, we used the global exposure mortality model (GEMM) to describe the relationship between pollution exposure and mortality. Compared to prior studies that apply integrated exposure-response functions (IER), our health cost estimates capture not only the 6 major causes (ischemic heart disease, stroke, chronic obstructive pulmonary disease, lung cancer, lower respiratory infections and type 2 diabetes) of deaths previously attributable to PM_2.5_ pollution, but also additional non-accidental causes of mortality impacts. This study provides updated estimates of the global economic cost of deaths attributable to ambient air pollution in 195 countries from 2000 to 2016. Furthermore, our decomposition analysis shows the key factors associated with the change in global health economic cost of air pollution over time. The findings of this study are particularly relevant for pollution control policies in countries that face both high levels of pollution and a rapidly-ageing population.

### Implications of all the available evidence

Air pollution’s impact on mortality disproportionately affects the older segments of the population. Given general trends in population aging, our results suggest that the economic health costs of ambient air pollution will continue to rise in the immediate future. Although air quality management is needed globally, it is especially urgent for highly-polluted and highly-aged countries. In addition, targeting health care and preventative interventions towards older individuals, such as by providing them with at-home air filtration systems, can partially mitigate the health costs of ambient air pollution. Understanding the health effects of air pollution across the age distribution has important implications for assessing the aggregate costs of other age-differential health risks, such as the outbreak of COVID-19. Since the outbreak is particularly dangerous for the elderly with preexisting health conditions, air pollution might exaggerate the health risks of the epidemic. The methodology developed in this study is applicable for evaluating the health economic costs of the epidemic. Further investigation on the indirect health cost of COVID-19 due to the previous exposure of air pollution might provide more incentives for air pollution control.

## II Methodology

### 1. Estimating global PM_2.5_ exposure from 2000–2016

We retrieved the global estimates of PM_2.5_ concentrations using a database that combines satellite products and ground-based measurements with chemical transport modelling, applying a geographically weighted regression (GWR) at 0.1° − 0.1° (~11 km at the equator) resolution.^17^ These estimates were more consistent with ground-based monitoring data than PM_2.5_ estimates without a GWR adjustment.

To estimate the population that was exposed within each grid of PM_2.5_ concentration, we collected population data (Gridded Population of the World - GPW v4) from NASA’s Socioeconomic Data and Applications Center (SEDAC), which is gridded at 0.0083° − 0.0083° (~1 km at the equator) resolution.^18^ We aggregated the population data into the same (0.1° × 0.1°) resolution as the PM_2.5_ data.

### 2. Estimating age-specific health risks from 2000–2016

We applied the GEMM to estimate the health risks from all non-accidental mortality due to exposure from PM_2.5_ pollution. We also estimated deaths associated with five specific causes—chronic obstructive pulmonary disease (COPD), ischemic heart disease (IHD), stroke, lung cancer (LC) and lower respiratory infection (LRI)—to understand their contribution to the total mortality.^19^ We defined non-accidental causes of mortality as all noncommunicable diseases (NCD) and LRI. We subtracted the mortality related to the five specific causes of disease (5COD) from all non-accidental causes of mortality (NCD + LRI) to compute the additional non-accidental mortality (NCD + LRI − 5COD). We classified age groups in 5-year increments, censoring the last age group at 85 years and older. The hazard ratio (here equated with relative risk, or RR) of each cause of death *k*, at age *m*, in grid cell *i*, and year *j* is represented by the following function:

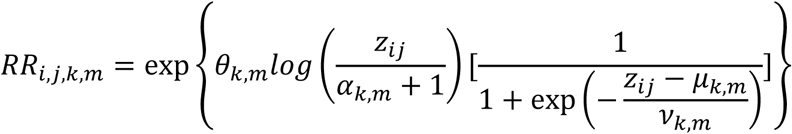

where *z_ij_ = C_ij_* − *cf, C_ij_* is the ambient concentration of PM_2.5_ in grid *i* and year *j*, and *cf* is the counterfactual concentration of PM_2.5_ below which there is assumed to be no additional risk. We assumed that the counterfactual concentration follows a uniform distribution between 2.4 μg/m^3^ and 5.9 μg/m^3^.^5^ The functional form of GEMM is developed from the transformation of a log-linear model. In the GEMM, *θ* and its SE control the slope of a non-linear regression that represents the relationship between exposure concentration and RR, and *α* defines the curvature of the model.^5^ Burnett et al. (2018) reported age-specific parameters of *θ, α, μ, ν* from 41 cohort studies from 16 countries.

### 3. PM_2.5_ related deaths and years of life lost (YLL) from 2000–2016

Previous studies have estimated both the number of lives lost and the shortened life expectancy due to long-term exposure to air pollution.^20–22^ Studies have discussed whether valuing lives or life-years lost is a more appropriate metric for estimating the health impacts attributable to air pollution. The WHO treats deaths and YLL as equally legitimate metrics for measuring the cost of air pollution.^23,24^ While Rabl (2003) argues that the number of deaths is not an appropriate metric for air pollution related mortality. As long-term exposure to air pollution contributes to deaths by shortening life expectancy,^26^ we computed PM_2.5_ related YLL as the main health metric for valuing health costs.

We estimated attributable deaths using the following equation:

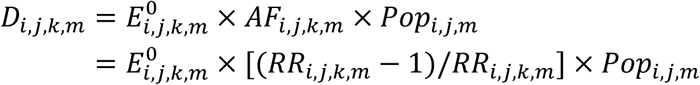

Given *D_i,j_,_k,m_* the YLL of each cause *k*, at age *m*, in grid cell *i*, and year *j* is derived as follows:

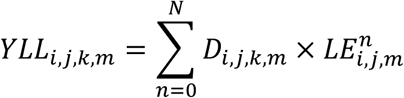

where *D_i,j,k,m_* is the number of prevalent deaths, 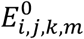 is the baseline mortality rate of the exposure population, *AF_i,j,k,m_* is the attributable fraction of relative risks caused by air pollution, *Popij_m_* refers to the exposed population, *YLL_i,j,k,m_* is the years of life lost, and 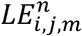 is the life expectancy. We estimated attributable deaths and YLL with cause-specific, location-specific, year-specific, and age-specific parameters. While attributable mortality and population are observable at the grid cell level, we obtained the baseline mortality of each cause and the life expectancy for each age group at the country level using data from the 2017 Global Burden of Disease (GBD) Study.^27^

### 4. Estimating age-adjusted VSL and VSLY from 2000–2016

This study explored the heterogeneity of health economic costs with respect to the age of the exposed population by adopting an age-adjusted VSLY. Whether VSL and VSLY vary with age has gained much attention in the literature.^28,29^ Most previous studies have adopted constant VSL or VSLY as a valuation measure to estimate health economic cost of air pollution.^30,31^ However, theoretical and empirical findings showed that VSL and VSLY can vary substantially with population age.^14,15,29^ Intuitively, the VSL decreases with age since the older population expect relatively fewer remaining life years. From this perspective, individuals’ willingness-to-pay (WTP) to avoid fatality risk would peak at birth (assuming that parents express WTP for avoiding deaths of their own children) and declines continuously afterwards. Following this assumption, the *Lancet* Commission on Investing in Health developed an age-adjusted value of standardized mortality unit, which assumes that the VSL is linearly correlated with one’s remaining life expectancy.^13^ However, this measure of VSL may not accurately reflect society’s preferences for avoiding mortality risk.^32^ Studies find that WTP for a unit of fatal risk reduction also varies with the quality of life year and wealth over a lifetime.^33,34^ This indicates that both VSL and VSLY change with health, longevity and wealth over the course of one’s lifetime. Consistent with this argument, studies that specifically examine the age-VSL relationship have found that it reflects an inverted-U shape.^14,15,35^

To account for the variation in WTP over the lifecycle, we developed an age-adjusted VSLY that modifies the constant VSL for wealth, remaining life expectancy, and age-specific survival rates at the country level. First, we adjust for the effect of wealth over the lifecycle by multiplying the constant VSL with age-specific wealth weights (wealth for certain age group divided by the mean wealth of all-age group) that capture changes in consumption over a lifetime. Individual wealth does not evolve monotonically with age.^36,37^ Resources and wealth tend to accumulate before retirement age and then decrease modestly after retirement.^38^ Figure 1a (see appendix pp 2) illustrates that the wealth ratio increases with age and decreases after ages 65–70, while the ratio of aged population is higher than the wealth of the population younger than 50 years old. We assumed that those under 18 years share the same wealth weight as their family members. This wealth-age profile suggests that older people, who have more wealth to lose if they die, are likely to have a higher willingness to pay to avoid a unit of healthy life year loss compared to younger adults with less to lose.^39^ Second, we adjusted the constant VSL by the ratio of the age-specific remaining life expectancy over the mean life expectancy of the total population. The country-specific constant VSL was estimated using a benefit-transfer approach assuming a base VSL of US$3.54 million estimated from OECD countries.^40,41^ This adjustment indicates that the VSL decreases with individuals’ remaining life years. Third, we adjusted the VSL by the survival probability to represent the quality of a life year. Individuals with a low survival probability have little incentive to spend money to avoid fatal mortality due to a low quality of life and few remaining life years.^39^

**Figure 1.**
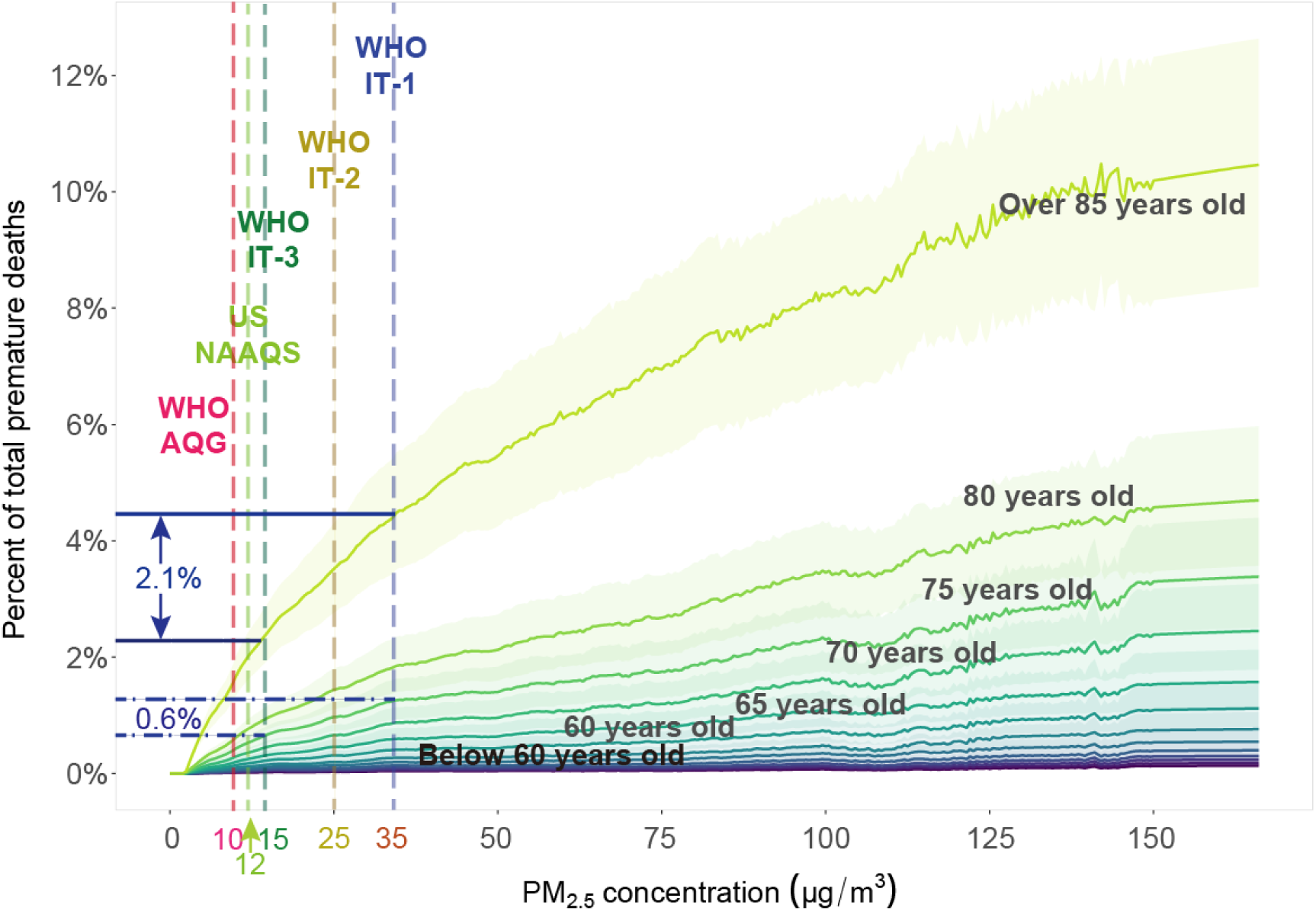
Percent of deaths attributable to PM_2.5_ out of population by age group over a range of PM_2.5_ concentrations in 2016. Shades around each line represent 95% of uncertainty intervals of each age group. The attributable mortality is estimated by age group over 350 bins of PM_2.5_ concentration. The noise in the lines of attributable mortality originated from the geographical variation in age-specific mortality.

The value of age-adjusted VSLY is calculated from a remaining life-expectancy- and wealth-adjusted VSL and survival rate by age:

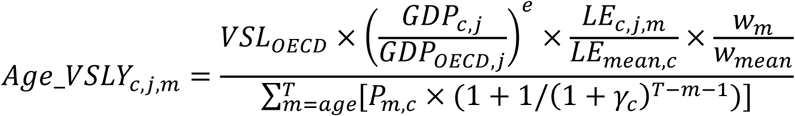

where *Age_VSLY* is the age-adjusted value of a statistical life year, *VSL_OECD_* is the base VSL from OECD countries, *GDP_c,j_* is the gross domestic product (GDP) per capita for country *c* in year *j*, *GDP*_*oecd,j*_ is the average GDP per capita of OECD countries, *e* is the elasticity of income in VSL, *w_m_* is the average wealth at age *m, w_mean_* is the average wealth, *T* is the age of expected death at age *m, P_m,c_* is the survival probability at age *m*, and *γ* is the discount rate. We defined 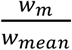 as the wealth weight. Country-specific GDP per capita from 2000 to 2016 was obtained from the World Bank database.^42^ GDP per capita, wealth, VSL and VSLY are adjusted to purchasing-power-parity (PPP) dollars in 2011 in each country. Details regarding the estimation of constant VSL and VSLY, and the age-adjusted VSL are provided in the appendix (pp 1).

The estimation procedure involved a range of parameters that have direct and indirect impacts on the economic cost estimates. According to previous studies, the income elasticity of VSL is about 1.0 for non-US countries.^43^ Similarly, a meta-analysis of global VSL studies reports that the income elasticity of VSL is between 0.9 and 1.3.^44^ Following the recommendations of the World Bank report, we selected a wide range of income elasticities for two categories of countries.^8^ For low- and middle-income countries, we select an income elasticity of 1.2, with a range of 1.0 to 1.4 for our sensitivity test. For high-income countries, we select an elasticity of 0.8 with a range of 0.6 and 1.0 for our sensitivity test. We set the discount rate at 6 percent for low- and middle-income countries, and 4 percent for high-income countries.^8^ The wealth weights are calculated based on age-specific wealth distribution data extracted from national statistical offices. Since the national wealth data of different age groups are very limited, we only obtained age-wealth distribution data for five countries: the United States, Canada, the United Kingdom, Germany and China.^45–50^ We then aggregated the country-level data into an global average wealth data by age. We ran a loess regression model to simulate a global average wealth weights in different age groups (appendix pp 1–2).

### 5. Valuation of the economic cost of air pollution

We estimated the economic cost within each age group as the product of the number of prevalent YLL (*Death* × *LE*) in that group and the discounted present value of a life year loss. In contrast to previous studies that used an age-invariant VSLY, we modelled the value of a statistical life year at each age by its age-adjusted VSLY.

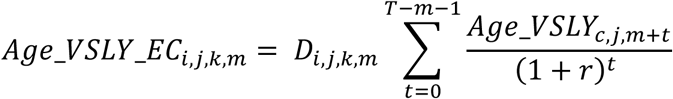

where *Age_VSLY_EC_i,j,k,m_* represents the economic cost of PM_2.5_ using the age-adjusted VSLY measure.

### 6. Uncertainty analysis and sensitivity test

Uncertainties in the distribution of PM_2.5_ concentration, exposed population sizes, life expectancy, exposure-response functions, socioeconomic parameters and valuation methods propagated to the health economic cost estimates. We adopted Monte Carlo simulations to estimate 95% uncertainty intervals (UIs) from 1000 draws of parameters and exposure-response functions in the health economic cost assessment.

For our sensitivity analysis, we evaluated the health cost using four additional valuation measures: i) a country-specific, constant VSLY across all age groups, ii) a global average age-adjusted VSLY, iii) a country-specific, age-adjusted value of statistical life (VSL), and iv) a country-specific, constant VSL across all age groups. To compare the health economic cost across countries and regions around the world, we also introduced a set of global average age-adjusted VSLY that removes differences due to variation in GDP per capita across countries. All simulations were done with R version 3.6.0.

## III Results

### 1. Disproportional health and economic cost on the elderly

Figure 1 illustrates the variation in age-specific mortality attributable to PM_2.5_ exposure. For each additional unit of PM_2.5_ concentration, the average attributable mortality of people aged 60 and older was 12 times (95% UI: 11, 13) higher than the risks of those under 60. While a decrease in ambient PM_2.5_ from 35 μg/m^3^ (WHO interim target 1) to 15 μg/m^3^ (WHO interim target 3) leads to a 2.1% decrease in the attributable mortality rate among those age 85 and older, this rate decreased by only 0.6% in those 75–80 and it decreased even less in younger ages. If PM_2.5_ was reduced to the WHO Air Quality Guideline (WHO AQG for PM_2.5_ is 10 μg/m^3^), the attributable mortality for younger ages ranged between 0.02% and 0.1%. However, the older population would still experience large risks, especially for those aged 70 and older (0.4% to 1.6%). These estimates suggest that the impact of PM_2.5_ pollution will grow substantially as the share of the population aged 60 and older expands. Therefore, the current WHO AQG, which is determined by the average effects of air pollution on general population without considering the effects of population ageing, might require strengthening. This suggestion is further augmented by several recent analyses showing that significant health impacts can still occur at levels of PM_2.5_ below the guideline.^51–53^

From 2000 to 2016, PM_2.5_ related health impacts increased significantly, particularly for the older segments of the population. Globally, 6.23 million (4.75 million to 7.79 million) deaths were attributable to PM_2.5_ in 2000, and the number grew to 8.42 million (6.50, 10.52) in 2016. The deaths in those aged 60 and older comprised more than 65% (66%-73%) of the total. Further, based on these estimates, deaths attributable to PM_2.5_ accounted for 12% to 15% of global all-cause deaths between 2000 and 2016. The deaths attributable to PM_2.5_ accounted for 138.49 million (94.76, 186.73) and 163.68 million (116.03, 219.44) YLL in 2000 and 2016, respectively (Table 1). The YLL of the older population accounted for 37% (30% - 40%) of all YLL attributable to PM_2.5_ pollution in 2016. Between 2000 and 2016, the YLL for those 60 years and older increased by 60%, while the YLL only increased by 3% among those younger than 60 years old.

Table 1 summarizes the number of deaths and YLL attributable to PM_2.5_ in the GBD super regions. The highest attributable mortality was 1.72 per 1000 persons (1.35, 2.10) in the *South Asia* super region followed by the *Southeast Asia, East Asia and Oceania* super region. The *Southeast Asia, East Asia and Oceania* super region was subject to the highest elderly YLL with more than 21.11 million (16.39, 26.27) YLL in the older population in 2016. Among all countries in these two GBD super regions, India and China were the largest contributors to these losses.

**Table 1.** Global health damages attributable to PM_2.5_ by GBD super regions in 2016.

| GBD super region | Population<br>(million) |  | Deaths<br>(million) |  | Years of life lost<br>(million) |  | Deaths per thousand |  | YLL per capita |  |
| --- | --- | --- | --- | --- | --- | --- | --- | --- | --- | --- |
|  | Elderly | Overall | Elderly | Overall | Elderly | Overall | Elderly | Overall | Elderly | Overall |
| **Global** | **898** | **7379** | **5.61**<br>**(4.46, 6.83)** | **8.42**<br>**(6.50, 10.52)** | **60.17**<br>**(46.54, 75.46)** | **163.68**<br>**(116.03, 219.44)** | **6.41**<br>**(5.09, 7.80)** | **1.17**<br>**(0.90, 1.46)** | **0.069**<br>**(0.053, 0.086)** | **0.004**<br>**(0.002, 0.006)** |
| Central Europe, Eastern Europe, and Central Asia | 78 | 415 | 0.47<br>(0.38, 0.56) | 0.63<br>(0.51, 0.75) | 4.53<br>(3.66, 5.42) | 9.38<br>(7.36, 11.53) | 6.16<br>(5.02, 7.30) | 1.55<br>(1.25, 1.85) | 0.059<br>(0.048, 0.071) | 0.02<br>(0.02, 0.03) |
| High-income | 246 | 1045 | 0.75<br>(0.61, 0.90) | 0.86<br>(0.69, 1.02) | 7.70<br>(6.16, 9.29) | 11.39<br>(9.11, 13.77) | 3.13<br>(2.54, 3.75) | 0.84<br>(0.68, 1.01) | 0.032<br>(0.026, 0.039) | 0.011<br>(0.009, 0.014) |
| Latin America and the Caribbean | 60 | 567 | 0.14<br>(0.11, 0.17) | 0.21<br>(0.16, 0.25) | 1.66<br>(1.30, 2.06) | 4.19<br>(3.24, 5.26) | 2.34<br>(1.87, 2.85) | 0.37<br>(0.30, 0.46) | 0.028<br>(0.022, 0.035) | 0.008<br>(0.006, 0.010) |
| North Africa and Middle East | 43 | 573 | 0.25<br>(0.19, 0.32) | 0.42<br>(0.30, 0.55) | 2.84<br>(2.05, 3.82) | 9.30<br>(6.12, 13.45) | 5.97<br>(4.52, 7.61) | 0.75<br>(0.54, 0.99) | 0.068<br>(0.049, 0.091) | 0.017<br>(0.011, 0.024) |
| South Asia | 147 | 1735 | 1.78<br>(1.43, 2.14) | 2.91<br>(2.28, 3.55) | 18.40<br>(14.40, 22.81) | 58.69<br>(42.84, 75.75) | 12.43<br>(9.99, 14.97) | 1.72<br>(1.35, 2.10) | 0.128<br>(0.101, 0.159) | 0.035<br>(0.025, 0.045) |
| Southeast Asia, East Asia, and Oceania | 277 | 2063 | 1.87<br>(1.49, 2.28) | 2.49<br>(1.96, 3.05) | 21.11<br>(16.39, 26.27) | 42.09<br>(31.95, 53.36) | 6.94<br>(5.52, 8.42) | 1.23<br>(0.98, 1.52) | 0.078<br>(0.061, 0.097) | 0.021<br>(0.016, 0.027) |
| Sub-Saharan Africa | 46 | 981 | 0.35<br>(0.25, 0.47) | 0.92<br>(0.59, 1.34) | 3.93<br>(2.57, 5.80) | 28.63<br>(15.40, 46.31) | 7.61<br>(5.45, 10.23) | 0.96<br>(0.61, 1.40) | 0.087<br>(0.057, 0.128) | 0.030<br>(0.016, 0.048) |

The health economic cost of the YLL attributable to PM_2.5_ pollution in the total population increased from US$2.37 trillion (1.88, 2.87) to US$4.09 trillion (3.19, 5.05) per year from 2000 to 2016, reflecting a 70% increase in this period. This cost represents 3.6% (2.8%, 4.5%) of global GDP in 2016. The health cost to the older population alone, however, increased from US$1.34 trillion (1.09, 1.60) in 2000 to US$2.40 trillion (1.89, 2.93) in 2016. The increase in economic cost over this period was 20% faster in the older population than in the younger population. Globally, the health economic cost per capita of the older population was 10.2 times (8.1, 12.5) the per capita cost of the younger population. Hence, increases in air pollution disproportionately increases the economic cost with respect to the older population.

Over the 17-year period, PM_2.5_ attributable economic cost in the older population accounted for 57% to 59% of that in the total population. Importantly, despite generally lower air pollution concentrations, the *High-income* super region had the highest share of total economic cost borne by the elderly population (from 67% to 72%), followed by the *Central Europe, Eastern Europe, and Central Asia* super regions (from 56% to 60%). In comparison, given its much younger age distribution, only 24% of the total cost in the *Sub-Saharan Africa* super region was borne by the older population. The share of the health economic cost on the older population increased over time in most countries due to rapid population ageing (Figures 2C & 2D). The proportion of economic cost related to the older population was higher in countries with relatively lower population-weighted PM_2.5_ concentrations as there was an overall negative correlation between PM_2.5_ levels and the national proportion of those older than 60 years (Appendix Figure 3a). For example, Japan experienced the highest proportion of the health economic cost (from 70% to 83%) in the elderly from 2000 to 2016, followed by Italy (from 76% to 81%), Spain (from 71% to 77%) and Germany (from 70% to 76%) where the population weighted PM_2.5_ concentration was under 20 μg/m^3^.

**Figure 2.**
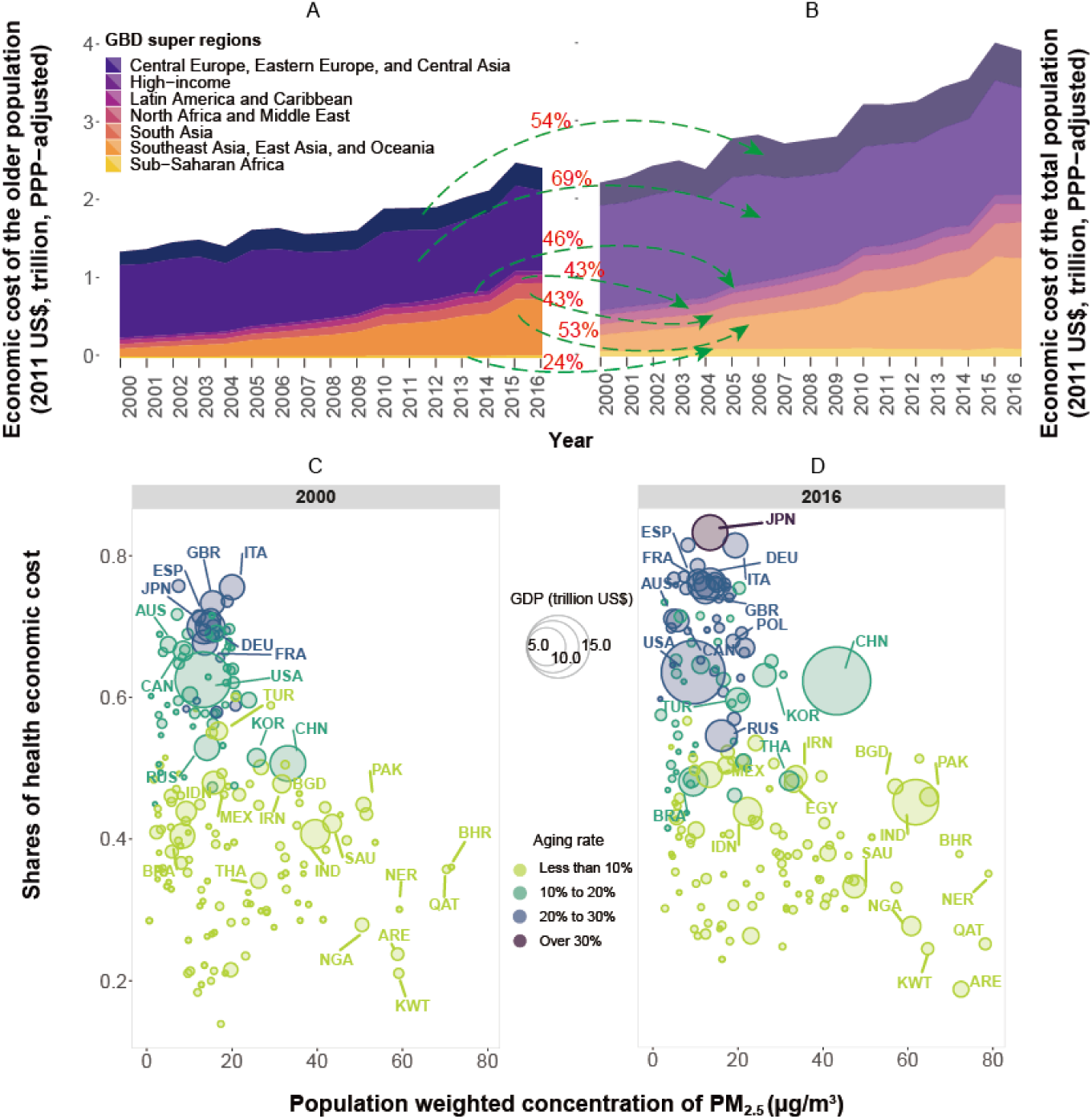
Shares of economic cost by GBD super regions and 195 countries and territories Figure 2A illustrates the health economic cost in people aged 60 and older from 2000 to 2016, and Figure 2B presents the economic cost in the total population. The values with the green arrows are the average shares of health economic cost with respect to the elderly population in each GBD super region over the 17-year period. Figures 2C and 2D show the growing share of the health economic cost in the older population from all-age health loss as a function of the population weighted concentration of PM_2.5_ in 195 countries and territories from 2000 to 2016. National GDPs are adjusted to purchasing power parity in 2011.

### 2. Economic cost comparison by age and cause

The health damage attributable to PM_2.5_ varied substantially by age and cause. Among all the age groups, those aged 85 and older had the highest the number of deaths. Excluding the over-85 and under-20 age groups, mortality counts peaked at age 70–75 years with 0.85 million [0.67, 1.04] deaths, and YLL was largest for the aged 60–65 population with 14.89 million (11.48, 18.77) years lost (Figure 3A & 3B). Between 2000 and 2016, deaths and YLL decreased in the population under 30. However, the health economic cost increased in this age group over the study period. The highest health economic costs were associated with the populations aged 60–65 (US$0.48 trillion [0.38, 0.58]) and the aggregated age group of people over 85 years old (US$0.53 trillion [0.42, 0.65]).

To account for differences in age structure between countries and to compare the health cost of PM_2.5_ across regions, we adopted a global average age-adjusted VSLY to estimate the cost of YLLs attributable to PM_2.5_ (Figure 3D). The results show that the largest health economic costs occurred in the *South Asia* super region *and Southeast Asia and East Asia* super region, which accounted for 58% (57%-60%) and 64% (63%-66%) of the global health economic costs in 2000 and 2016, respectively.

**Figure 3.**
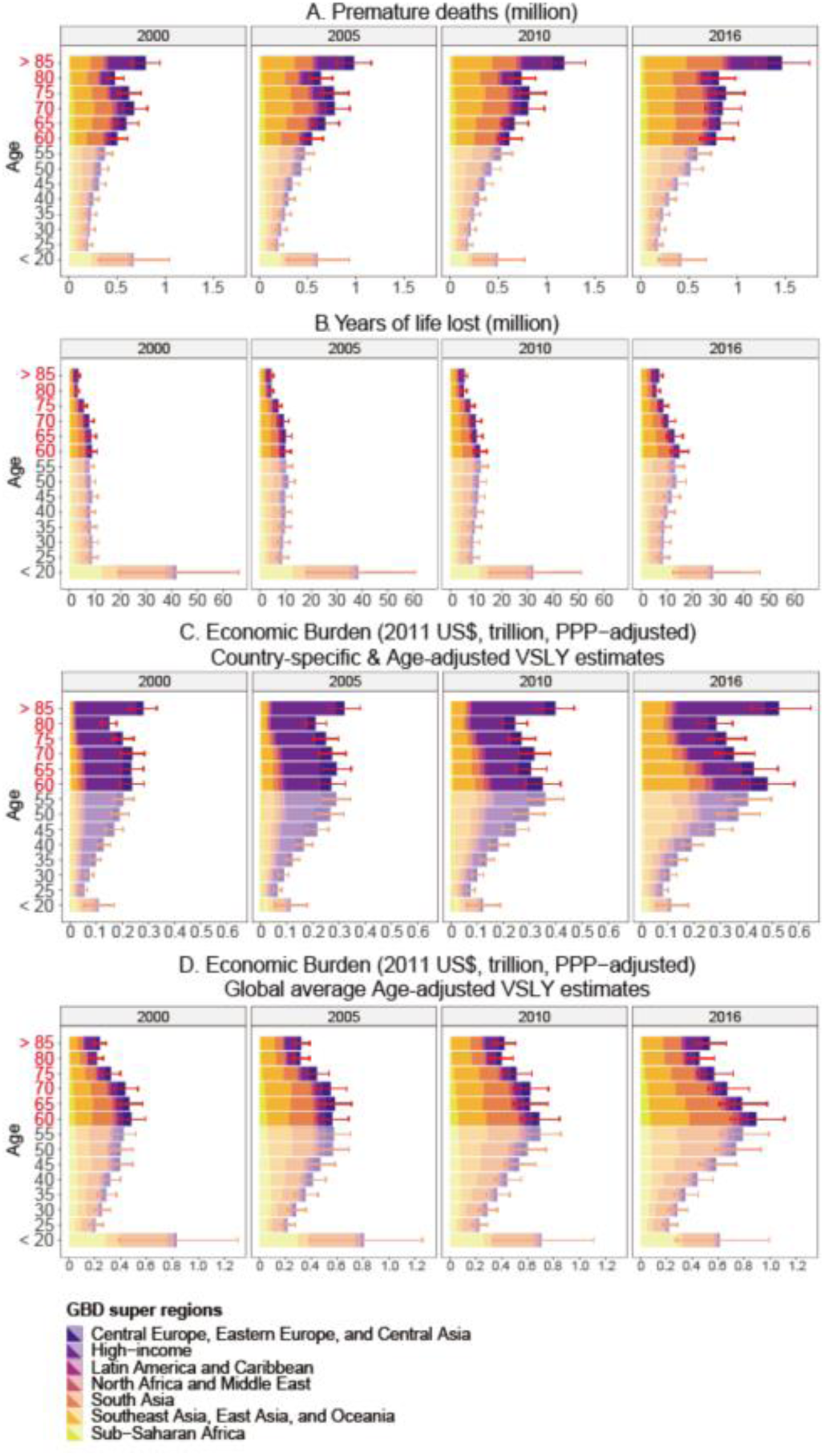
Attributable deaths, YLL and economic cost by age across the GBD super regions between 2000 and 2016. Figure 3A illustrates attributable deaths by age across regions; Figure 3B represents the corresponding YLL in each age group and region; Figure 3C illustrates the variation in health economic cost by age and region over time using a country-specific and age-adjusted measure of VSLY; Figure 3D shows the distribution of health economic costs using a globally averaged, age-adjusted VSLY. Red error bars represent 95% uncertainty intervals.

Globally, attributable deaths and YLL associated with additional (beyond the 5 diseases included in the 2016 GBD study) nonaccidental causes (NCD *+* LRI − 5COD) accounted for 30% of attributable deaths and YLL from PM_2.5_.^54^ Figure 4 indicates that additional non-accidental mortality comprised 44% (36%-56%) of the total health cost attributable to PM_2.5_, though this varied widely by country. At the national level, the health economic cost of additional non-accidental mortality comprised over 30% of the total loss attributable to PM_2.5_ pollution in China and India in 2016. In comparison, the share of cost due to additional non-accidental mortality in Japan and the United States was 56% (53%-64%). Overall, the North Africa, Asia and Central Europe regions had 30% of their total cost due to the additional non-accidental mortality, while the High-income and the Latin America regions had more than 50% of their health cost caused by additional non-accidental deaths. From 2000 to 2016, high-income countries generally experienced a decrease in health economic costs due to the five specific causes, but an increase from the additional non-accidental mortality. On the other hand, low-middle-income countries had faster growth in economic costs associated with the five specific causes than that due to additional non-accidental mortality.

**Figure 4.**
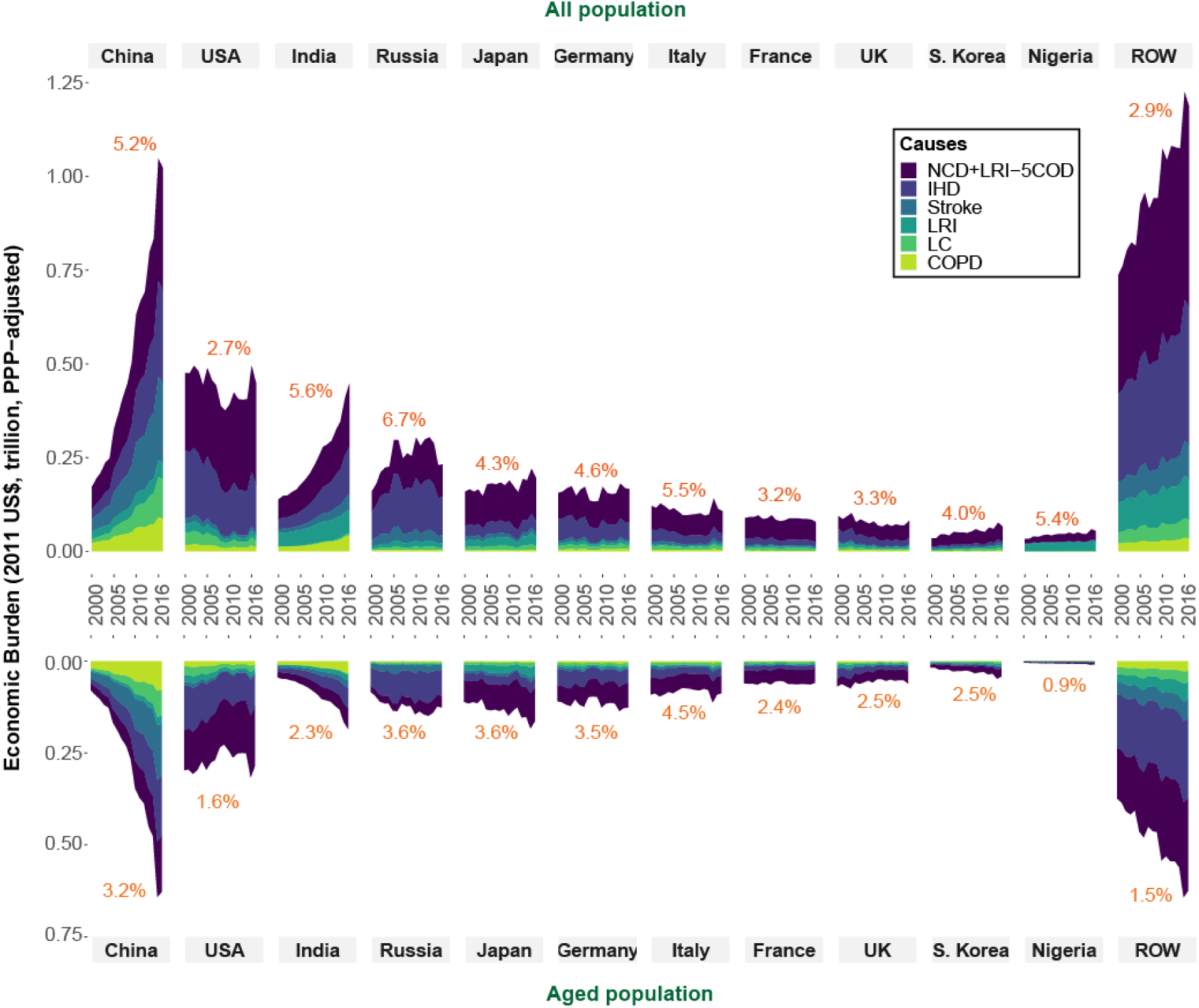
Selected countries and the rest of the world (ROW) economic cost of various causes related death from 2000 to 2016. The percentage in the graph represents the fraction of GDP in 2016. COPD is chronic obstructive pulmonary disease, IHD is ischemic heart disease, LC is lung cancer, LRI is lower respiratory infection, and (NCD + LRI − 5COD) is the additional non-accidental mortality.

The health economic costs of PM_2.5_ varied significantly among the five specific causes. IHD was associated with the highest costs among the five specific causes of disease, comprising 27% (26%-28%) of the total health economic cost attributable to PM_2.5_ in 2016. The cost of IHD was 5 to 10 times that due to the other four specific diseases. The global health costs associated with IHD increased from US$0.7 trillion (0.6-0.9) to US$1.1 trillion (0.9-1.4) from 2000 to 2016. The health cost associated with stroke increased fastest among all the specific causes, followed by COPD. The health economic cost of stroke in 2016 was 2.4 times that in 2000. More than 80% of the economic cost attributable to COPD was associated with the older population. This increased to an over 90% share in the aged countries, such as Japan, Italy, and Sweden. In contrast, countries such as the United Arab Emirates and Kuwait only had 30% of the economic costs caused by COPD related to the older population. In those older than 60 years, IHD, LC and stroke accounted for 63%, 68% and 64% of the total health economic cost attributable to PM_2.5_, respectively, in 2016.

Figure 4 also reports the percentage of GDP loss due to the health economic costs related to PM_2.5_ pollution in 2016. China, USA, India, Russia and Japan were the countries that had the highest health economic costs worldwide. However, the health economic cost relative to national GDP was higher in Russia, India, Italy, Nigeria and China.

### 3. Driving factors of growing health economic cost

The substantial health cost of air pollution reflects the influence of environmental, demographic and socioeconomic factors. We decomposed the change in health cost attributable to ambient PM_2.5_ pollution from 2000 to 2016 by country and region into five major contributors—population growth, population ageing, age-specific mortality, the exposure level of ambient PM_2.5_ and the growth of GDP per capita (see appendix for detail method, pp 2–3). Globally, the effects of population ageing offset 52% of the benefits gained from mortality reduction over the 17-year period. Figure 5 shows the relative and absolute contribution of the five driving factors of the rise in the health cost of ambient PM_2.5_ in the total population (Figure 5A) and the older population (Figure 5B) in the GBD super regions. Among all driving factors, the increase in GDP per capita was the dominant contributor to the rapid growth of health cost over the study period in the South-East Asian regions (Figure 5A-5B). In the *High-income* super region, the change in age-specific mortality played a key role in reducing the health cost, lowering it by 28.5% between 2000 and 2016 (Figure 5A). However, the growth of health cost due to population ageing offset 90% of the benefits of avoided deaths due to the reduction of mortality in the *High-income* super region (Figure 5A). Except for the South Asia and Sub-Saharan Africa super regions, all the other regions experienced 18.8% to 33.1% growth in health cost due to population ageing (Figure 5A).

**Figure 5.**
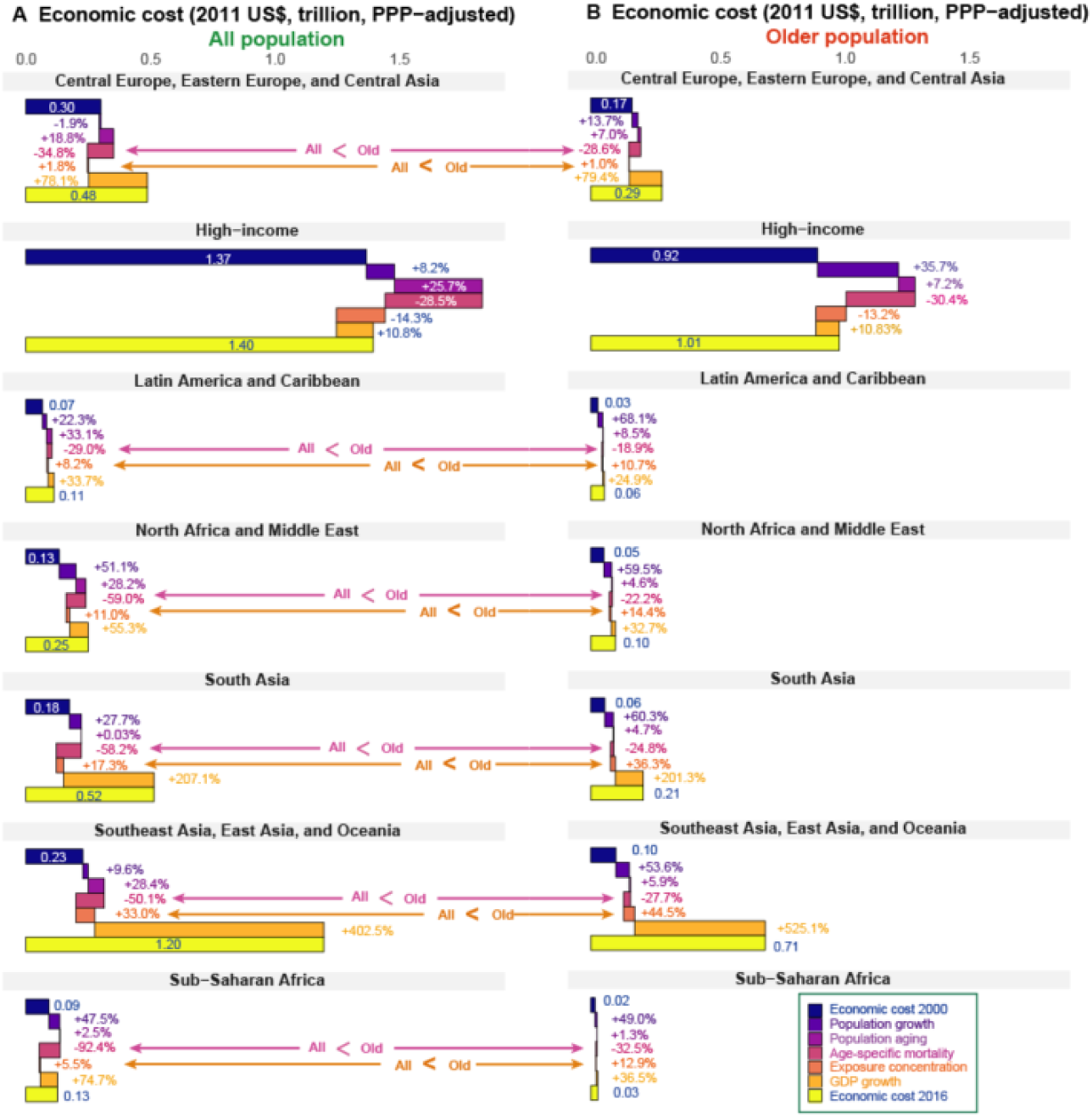
Contribution of each driving factor to the change of health cost related to ambient PM_2.5_ in the period of 2000–2016 in GBD super regions. The signs of “−” and “+” represent the negative and positive percent change in health cost. The first bars and the last bars are the total health cost in each GBD super region in 2000 and 2016. The length of each bar in between starts where the previous bar ends and represents the contribution of each factor between 2000 and 2016. The left graph (Figure 5A) is the decomposition of the driving factors for the total population and the right one (Figure 5B) is for the older population.

In terms of the older population, the contribution of population growth, population ageing, age-specific mortality and exposure of PM_2.5_ varied substantially (Figure 5B). The decrease in PM_2.5_ exposure benefited the older population more than the younger population. The effect of mortality reduction over this period avoided 56% of the health cost on the population under 60, while this reduction in mortality only brought a 29% decrease in health cost on the older population.

### 4. Comparative analysis of health economic costs

The health economic cost of air pollution changes significantly with the valuation methods on which the estimates are based. Therefore, as sensitivity analyses, we compared the health cost estimates computed using the country-specific age-adjusted VSLY with those using: 1) a country-specific constant (age-invariant) VSLY, 2) a global average age-adjusted VSLY, 3) a country-specific age-adjusted VSL, and 4) a country-specific constant VSL, respectively. Unlike the country-specific methods, the global average age-adjusted VSLY does not place a higher value on the lives of people from richer countries when estimating the global health costs, we used a global average age-adjusted VSLY measure to compare the health cost at the global level.

Estimates of health cost were highest using the constant VSL measure compared to those using the other four valuation measures. In 2016, the health economic cost was US$8.32 trillion (6.62, 10.11) using an age-invariant VSL, which was more than two times the estimates based on an age-adjusted VSL (US$3.88 trillion [3.07, 4.73]). In contrast, health costs were US$4.54 trillion (3.58, 5.57) and US$4.09 trillion (3.19, 5.05) based on an age-invariant VSLY and an age-adjusted VSLY, respectively. The global average age-adjusted VSLY indicated that the global health economic cost was US$5.09 trillion (4.06, 6.19) in 2016, which was higher than the estimate from the country-specific age-adjusted VSLY.

Globally, the health economic costs were unequally distributed across various levels of exposure to PM_2.5_ concentration. This substantial variation was mainly due to differences in GDP per capita and annual PM_2.5_ concentration across countries worldwide. In 2016, the global health economic cost estimated by country-specific valuation measures (i.e., constant VSL, age-adjusted VSL, constant VSLY and age-adjusted VSLY) peaked at regions with PM_2.5_ concentrations of 14.5 μg/m^3^ (Figures 6A, 6B, 6D, 6E). Applying these four valuation measures, we also found that 50% of global health economic cost occurred in regions where PM_2.5_ was below 25 μg/m^3^. In contrast, if a global average age-adjusted VSLY measure is applied, half of the all-age health economic cost occurred in regions where ambient PM_2.5_ was below 41.5 μg/m^3^. For the constant VSL, the share of economic cost among the ageing population was 77%, while the share dropped to 60% when the age-adjusted VSL was used. In comparison, the older population’s share of economic costs was 61% and 59% when we applied an age-invariant VSLY and an age-specific VSLY, respectively. Figure 5a (in appendix pp 6) shows that the United States, Japan and Russia, where a VSL was relatively higher due to the higher GDP per capita, had the highest health cost among the regions with PM_2.5_ levels below 25 μg/m^3^. In contrast, China and India had the highest health costs in the regions where annual PM_2.5_ was 25 μg/m^3^ to 125 μg/m^3^, whereas these countries had a relatively lower VSL.

**Figure 6.**
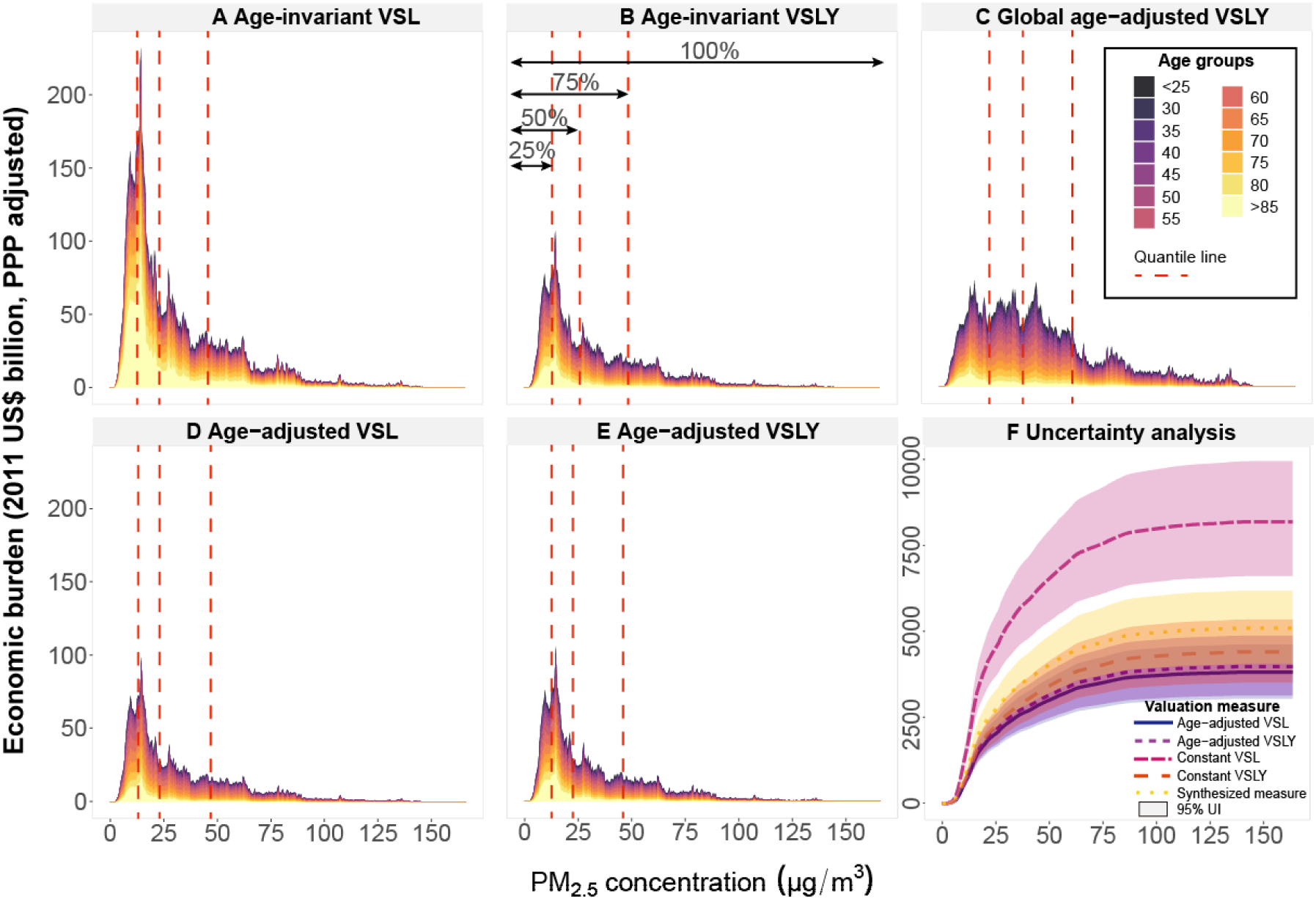
Global health economic cost distribution as a function of PM_2.5_ concentration by age group in 2016. Figures 6A-6E are area plots of the global age-specific health economic cost with the region level PM_2.5_ concentration. The integral of the area in each color represents the total health cost by age over a range of PM_2.5_ concentrations. Figure 6F illustrates the cumulative health economic costs with 95% UI, using four valuation measures and a synthesized valuation measure.

Figure 6F presents the 95% confidence intervals of four country-specific valuation measures and a synthesized measure. For the synthesized measure, we aggregated the costs and uncertainties of these four measures by assuming that each valuation measure has an equal weight and value for policymaking. The health economic cost synthesized from the four country-specific valuation measures was US$5.2 trillion (4.1, 6.2), which was 27% higher than the estimates using the age-adjusted VSLY and 60% lower than the loss based on an age-invariant VSL.

## IV Discussion

In this study, we examined the health economic cost of deaths by looking at the interaction between air pollution and global ageing. The population aged 60 years and older, which accounts for 10% to 12% of the global population, suffered 57%-59% of the total health economic cost of deaths attributable to PM_2.5_ over the 2000–2016 period, due to the combination of global ageing and the large impacts of air pollution on the older population. The health cost attributable to PM_2.5_ amongst this population was equivalent to 76% of the health spending in this age category in 2016,^55^ which leads to aggravating challenge to national healthcare systems. The rate of increase in the economic cost in the older population was 20% faster than that in the younger population. The disproportional health costs with respect to the older people varied across regions and countries. The fraction of economic costs on the population aged 60 years and older was highest in the *High-income* super region and lowest in the *Sub-Saharan Africa* super region.

The rapid growth of global economic cost was mainly driven by the increase in GDP per capita, population ageing and population growth over the period of 2000-2016. The effects of population ageing offset 52% of the benefits attributable to the overall reduction in global mortality. In addition, the benefits of mortality reduction did not distribute evenly across all age groups. Reduction in mortality contributed to a faster rate of health economic cost abatement in the younger population, which might enlarge the disproportional economic cost on the older population. The growth in global health economic costs increased most rapidly in the *Southeast Asia, East Asia and Oceania* super region. This increase in health cost was mainly due to the growth of GDP per capita, increasing exposure of PM_2.5_ concentration and population ageing. In comparison, the increase of health cost in the *High-income* super region was mainly due to population ageing and growing GDP per capita.

The health economic costs attributable to ambient PM_2.5_ also varied significantly by cause. In addition to the five previously considered specific causes of disease in the 2016 GBD, the health impacts of additional non-accidental causes related to PM_2.5_ increased rapidly. Among the five traditional specific causes, IHD resulted in the highest per capita economic cost, five times that caused by LRI. Per capita health costs associated with lung cancer in the older population showed the highest average growth rate compared with that of other specific causes of disease. In comparison, the younger population had the highest growth rate in per capita economic cost associated with COPD. The differences of health economic costs by cause provide additional information to policymakers to facilitate the allocation of public medical resources. The additional costs of non-accidental causes of YLL suggest that the economic benefits of pollution control lead to much greater health benefits than previous estimates.

The health economic cost of ambient PM_2.5_ pollution fluctuates significantly according to the valuation measure used. The constant VSL measure generated the highest estimate of economic cost in 2016 (US$8.32 trillion [6.6, 10.1]) compared to the other four methods (ranging from US$3.07 trillion to US$5.57 trillion). Compared with an age-invariant VSL, which is used in most previous studies, the health economic cost of PM_2.5_ pollution using an age-adjusted VSLY measure considers three major effects that influence VSLY over one’s lifetime: the change of remaining life expectancy, life quality and wealth weights. After the adjustment of VSLY by age, we obtained a set of VSL estimates that varied with age in an inverted-U shape that peaks at 40–50 years old (appendix pp 11), which is consistent with previous studies. Shepard and Zeckhauser (1984) illustrated in a “Robinson Crusoe” analysis that VSL increases with age among young adults, peaks at age 40, and then continuously drops when they get older. Simulation models of self-protection decisions that examine life-cycle consumption under age-specific mortality and dynamic wealth constraints also generated similar findings.^57^ The life-cycle model of VSL depends on imperfect markets in which an individual is not allowed to borrow or lend money over his/her life time to avoid moral hazard problems.^58^ Based on the hedonic model, Aldy and Viscusi (2007) estimated VSL peaks at age 46 years old, making an inverted-U shape, by observing market equilibria of tradeoffs between wages and fatal risks. Although there are concerns about the inequality caused by adjusting VSL by age, social preferences revealed from consumers’ choices and workers’ wage rates imply that both VSL and VSLY peak at middle age and decline afterwards.^15^ Therefore, an age-invariant VSL will lead to higher health economic cost estimates for the older population and an age-invariant VSLY will generate higher cost estimates for young people.^59^ Since air pollution disproportionately affects the older population, individuals’ risk-money tradeoff varies as their age, mortality rate and socioeconomic status change.^60^ Applying the age-adjusted VSLY improves our knowledge on the health cost of mortality attributable to air pollution, especially in an ageing society. The health economic cost estimated using the age-adjusted VSLY facilitates the optimal allocation of pollution control resources and public healthcare investment for populations of different ages.

Despite the advancements to estimating the health cost of air pollution made in this study, limitations remain. First, we applied methods that require a series of global and historical data inputs, which are measured with uncertainty. Although the satellite-based estimates are generally somewhat lower than the ground monitoring data, the PM_2.5_ estimates used in this study show a highly consistency (*R^2^=0.81*) with ground measurements.^61^ In addition, uncertainties in the population spatial distribution typically increase by the disaggregation of the age categories. Second, the exposure-response functions applied in this study also introduced uncertainties. Our estimates of economic costs caused by NCD+LRI were 40% higher than the total of five specific diseases in the older population using the GEMM. Due to the limited number of available cohort studies, the GEMM assumes that the prevalence of deaths for the additional causes, such as chronic kidney disease and dementia,^62,63^ is similar in all countries compared to the countries that are included in the 41 cohort studies. Including more cohort studies from highly-polluted countries and additional exposure-response functions for other NCDs in the GEMM will reduce the uncertainties in the estimates of deaths attributable to PM_2.5_ pollution. We also suggest that the exposure-response relationships of other NCDs should be further investigated to improve the estimates of health cost in additional NCDs. For example, the 2017 GBD Study included type 2 diabetes as another specific cause of disease, while the 2019 GBD study incorporated the estimates of deaths mediated by the impact of air pollution on birthweight and short gestation. Adding additional causes of disease related to PM_2.5_ might reduce the share of the health impacts of the older population relative to the total impact over all ages. In addition, as the GEMM is based on adult cohort mortality analyses, it may underestimate the overall impact of air pollution and specifically impacts on infant mortality and morbidity impacts on younger age groups.^64^ Third, uncertainties embedded in the age-adjusted VSLY. We translated the VSL or VSLY of all countries worldwide based on the VSL database of OECD countries, because national studies are limited. However, this database might underestimate the VSL in the United States and even other higher-income countries. For example, Viscusi (2018) estimated that the VSL in the US is US$10 million based on a hedonic wage approach, which is much higher than the benefit-transfer estimate (US$ 4.5 million in the US) used in this study. If we adopt a VSL of US$ 10 million in the US, the health cost of PM_2.5_ in the US will be US$1.02 trillion (0.81, 1.22). Estimates of the income elasticity for VSL also varies across countries. Therefore, a benefit-transfer method induces uncertainties in VSL and VSLY estimates in countries. In this study, we applied an age-invariant value of income elasticity within two groups of countries: low-income and high-income countries. To limit these uncertainties, we considered a wide range of elasticities between 0.6 and 1.4, which covers the estimates of elasticity reported in most previous studies.^40,44^ We acknowledge that elasticities are different across countries, but we believe our uncertainty assessment captured most of the bias caused by the variation of income elasticity in the VSL estimates. To adjust VSLY by age, we adopted a global average wealth weights by age due to limited statistical data at national level. More country-level wealth data by age will improve the robustness of the estimates in this study.

In conclusion, the health economic cost of deaths attributable to ambient PM_2.5_ among the older population accounted for a major share of total cost and has increased substantially from 2000 to 2016 owing to growing GDP per capita, population ageing and population growth. The health economic cost estimated by the age-adjusted VSLY measure provides further information for national and local government to design pollution control strategies and allocate healthcare resources by populations’ ages. Additional non-communicable disease, not previously considered, represent a large share of the total health economic costs, thus the benefits of pollution reduction might exceed previous estimates. If substantial pollution reduction, especially in highly-polluted and highly-aged countries, is not achieved, ambient air pollution will lead to rapid increases in health economic cost related to mortality, which might induce substantial burden on national healthcare systems. In addition, improved health care targeted towards older individuals and providing opportunities to reduce exposures can be helpful to reduce the health cost of ambient air pollution. Given that many of the measures that will protect people from air pollution are the same ones necessary to decrease greenhouse gas emissions, this study provides a rationale for measures that will improve health across generations through both improved air quality and reduced health impacts related to climate change as the century progresses.

## Data Availability

Requests for access to the data should be made to the corresponding author at. Data could be made available provided the applicant has appropriate ethics approval and approval from the authors.

## Acknowledgements

H.Y. acknowledges funding from the National Natural Science Foundation of China (grant 71904104) and the China Postdoctoral Science Foundation (grant 2019M650726). Z.L. acknowledges funding from Qiushi Foundation, the Resnick Sustainability Institute at California Institute of Technology and the National Natural Science Foundation of China (grant 71874097 and 41921005).

## Supplementary appendix

### 1. Country-specific age-invariant VSL and VSL estimation

In the absence of empirical estimates of country-specific VSL, this study derived age-invariant VSL by country using a benefit-transfer method based on meta-analysis for OECD countries ^1^:

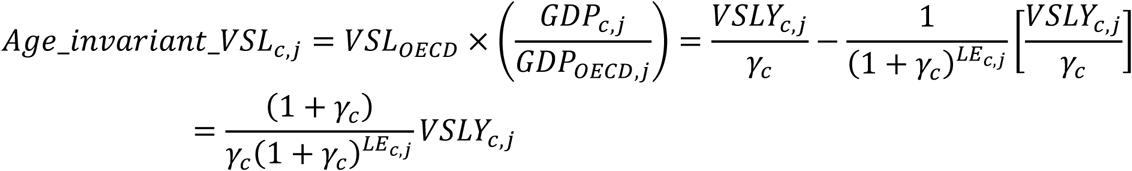

where *c* represents country, *j* is the year.

The discounted age-invariant VSLY is derived from dividing VSL by the remaining life years:

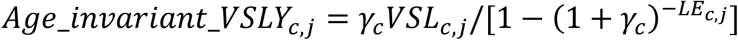

where *l* denotes individuals’ life span, *γ* is the discount rate, *VSL* represents the value of a statistical life, and *VSLY* is value of a statistical life year.

### 2. Country-specific age-adjusted VSL estimation

In this study, we generated five sets of economic cost estimates using four country-specific measures including age-adjusted VSL, age-adjusted VSLY, age-invariant VSL, age-invariant VSLY and one global average age-adjusted VSLY measures.

For loss of premature deaths, economic cost can be estimated using the loss of premature deaths multiplied by age-adjusted VSL or age-invariant VSL.

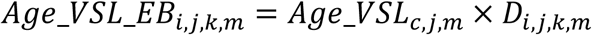

where *Age_VSL_EB_i,j,k,m_* represents economic cost of PM_2.5_ on the aged population using age-adjusted VSL measure.

### 3. Wealth weights estimation

Value of a statistical life is measured by a trade off between wealth and mortality rate (1-survival probability). Both wealth and mortality rates vary with age according to previous literature. To estimate the wealth distribution with ages, we obtained country-specific wealth database from USA, Canada, UK, Germany and China. To estimate country-specific and global average wealth weights, we adopted a Loess regression model to predict the wealth weight with respect to age. The wealth peaked around 65–70 years old in the United States while in China the wealth was highest among population aged 40–50. Due to the limited database for the wealth distribution with respect to age, we adopted global average wealth ratios by age aggregated from five countries. We ran a loess regression model to simulate a global average wealth weights in different age groups.

The detail of loess regression model is explained as following quadratic approximation:

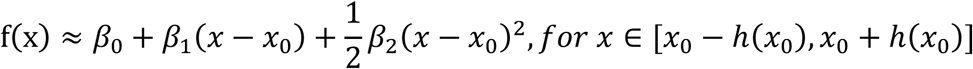

To estimate the local regression value 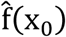, we solve the following equation to calculate the minimal β = (*β_0_, β*_1_*, β*_2_)′:

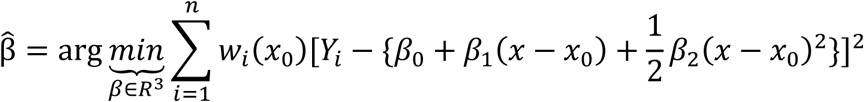

where 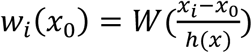, *h*(*x*_0_) is the span, here we applied a span of 0.3.

Figure 1a presents the loess regression results. Although the wealth of people at old age decreased slightly, it remains to be higher than most of the younger age population.

**Figure 1a.**
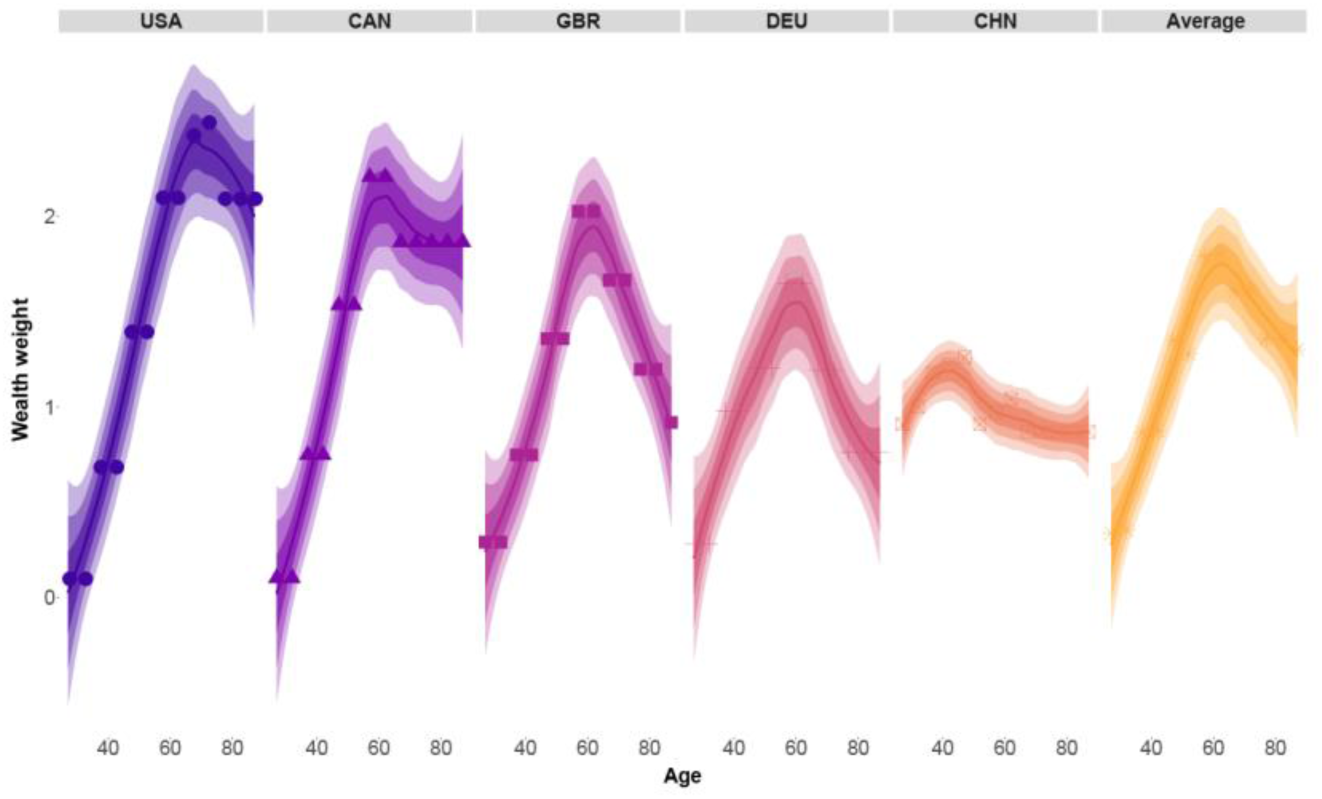
The wealth weights distribution with age

### 4. Driving factors decomposition

We decomposed five major driving factors—population growth, population ageing, age-specific mortality, exposure concentration and GDP per capita growth—related to health economic cost attributable to ambient PM_2.5_. According to GBD 2015 and Cohen et al. (2017),^2,3^ we simulated the attributable effects of each factor related to the change of health economic cost between 2000 and 2016. The baseline age-specific mortality effect was unrelated to the ambient PM_2.5_ exposure. All the analyses was done at four levels: global level, GBD super region level, income group level and country level.

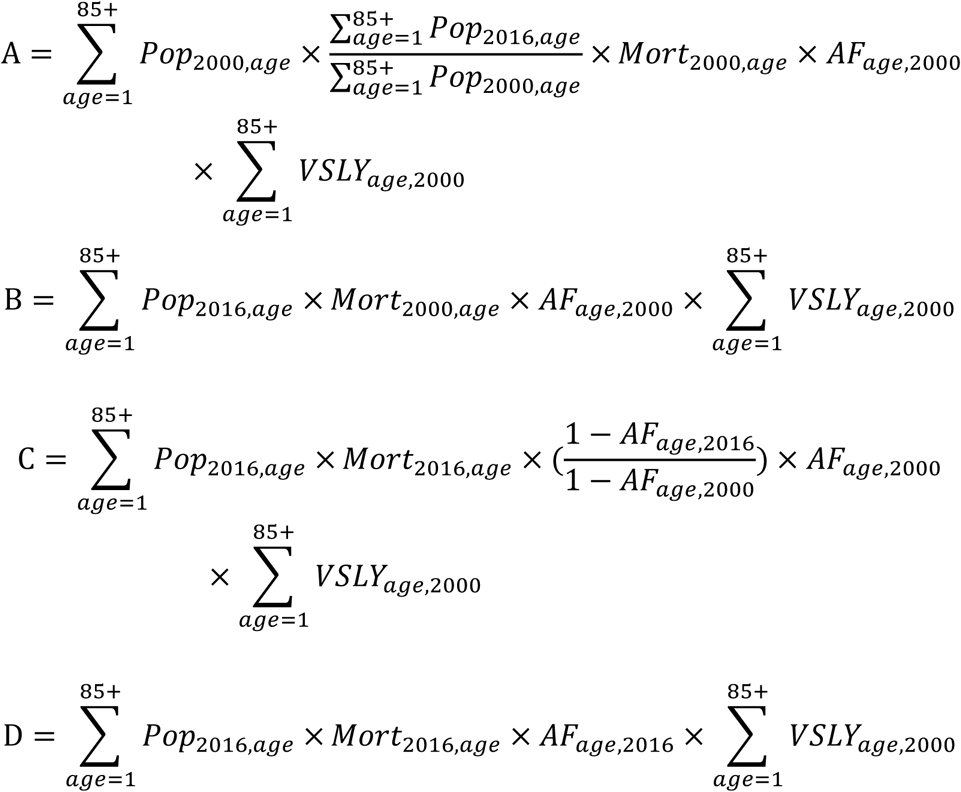

Therefore, the effects of five driving factors are estimated as follows:

Population growth effect (%) = (A − Age_VSLY_HC_2000_)/ Age_VSLY_HC_2000_

Population ageing effect (%) *=* (B − A)/Age_VSLY_HC_2000_

Baseline mortality change effect (%) = (C − B)/ Age_VSLY_HC_2000_

Exposure concentration change effect (%) = (D − C)/ Age_VSLY_HC_2000_

GDP growth effect (%) = ( Age_VSLY_HC_2016_ − D)/ Age_VSLY_HC_2000_

### 5. Health cost per capita in 195 countries

Figure 2a illustrates the health economic burden per capita between 2000 and 2016 by country for the aged population and all-age population. The health economic burden per capita of the aged population was much higher than the per capita loss of all-age population, both of which increased from 2000 and 2016.

**Figure 2a.**
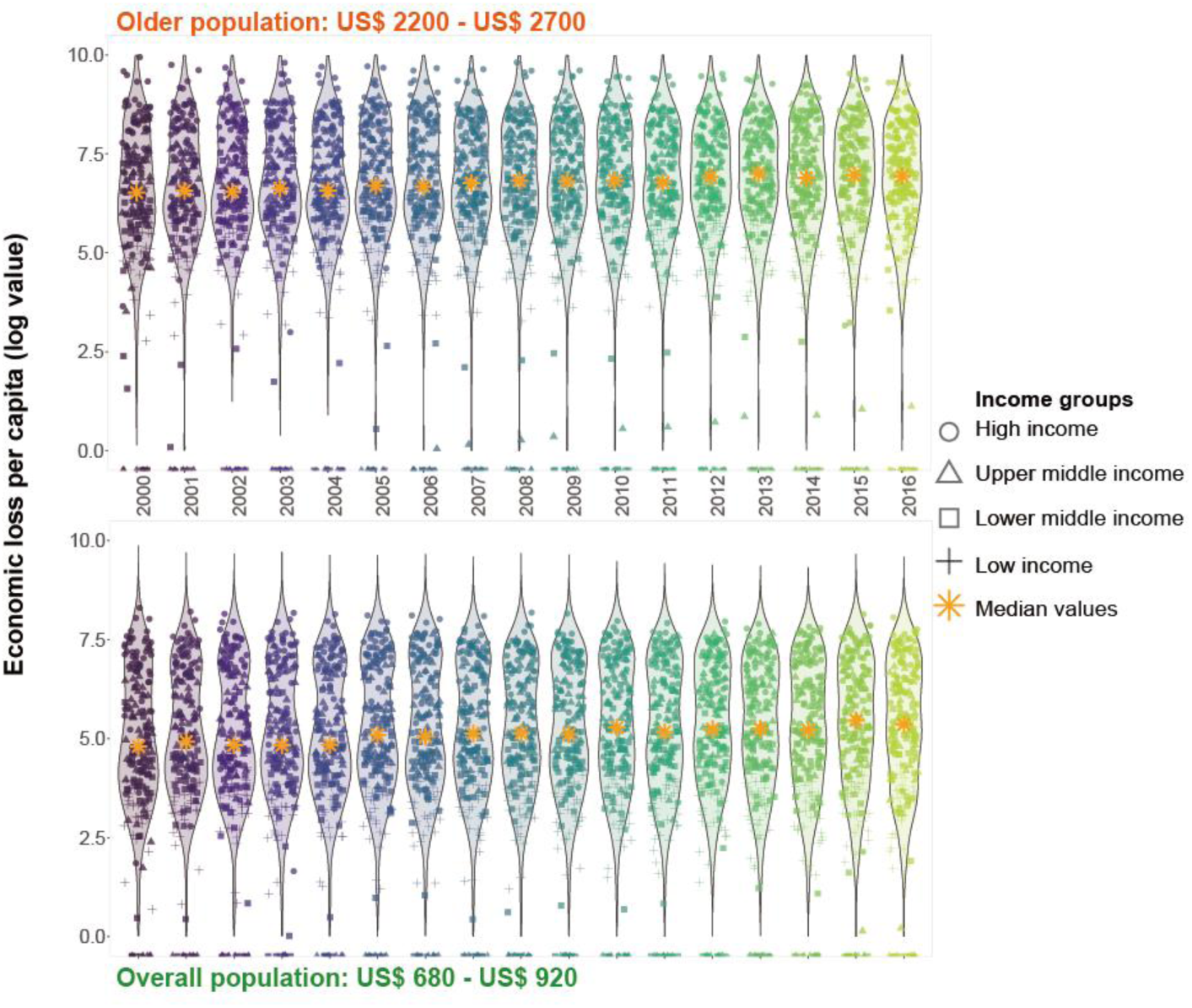
Health economic cost per capita by country in different income groups

### 6. Correlation between population ageing and PM_2.5_ concentration

We regressed the population ageing rates (proportion of population aged 60 years and older over the total population) with population-weighted PM_2.5_ concentration using a generalized additive model. Figure 3 a shows that the ageing rates decreased continuously with an increase of population-weighed PM_2.5_ concentration, which represents an overall decreasing trend of population ageing with an incremental population weighted PM_2.5_ concentration.

**Figure 3a.**
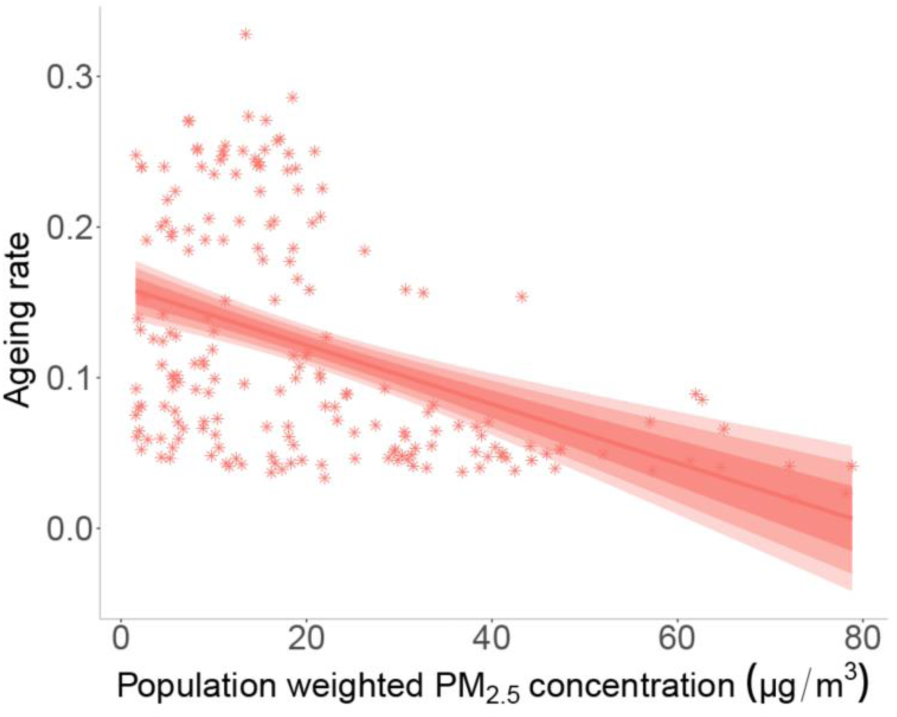
Correlation between population ageing and population weighted PM_2.5_ concentration in 2016. Shadows represent the 75%, 95% confidence intervals of predictions.

### 7. Health economic growth from 2000 to 2016

The health economic burden change of the aged population varied geographically between 2000 and 2016. Figure 4a shows that the highest economic burden growth happened in the East of China, India and the Central and Eastern Europe.

**Figure 4a.**
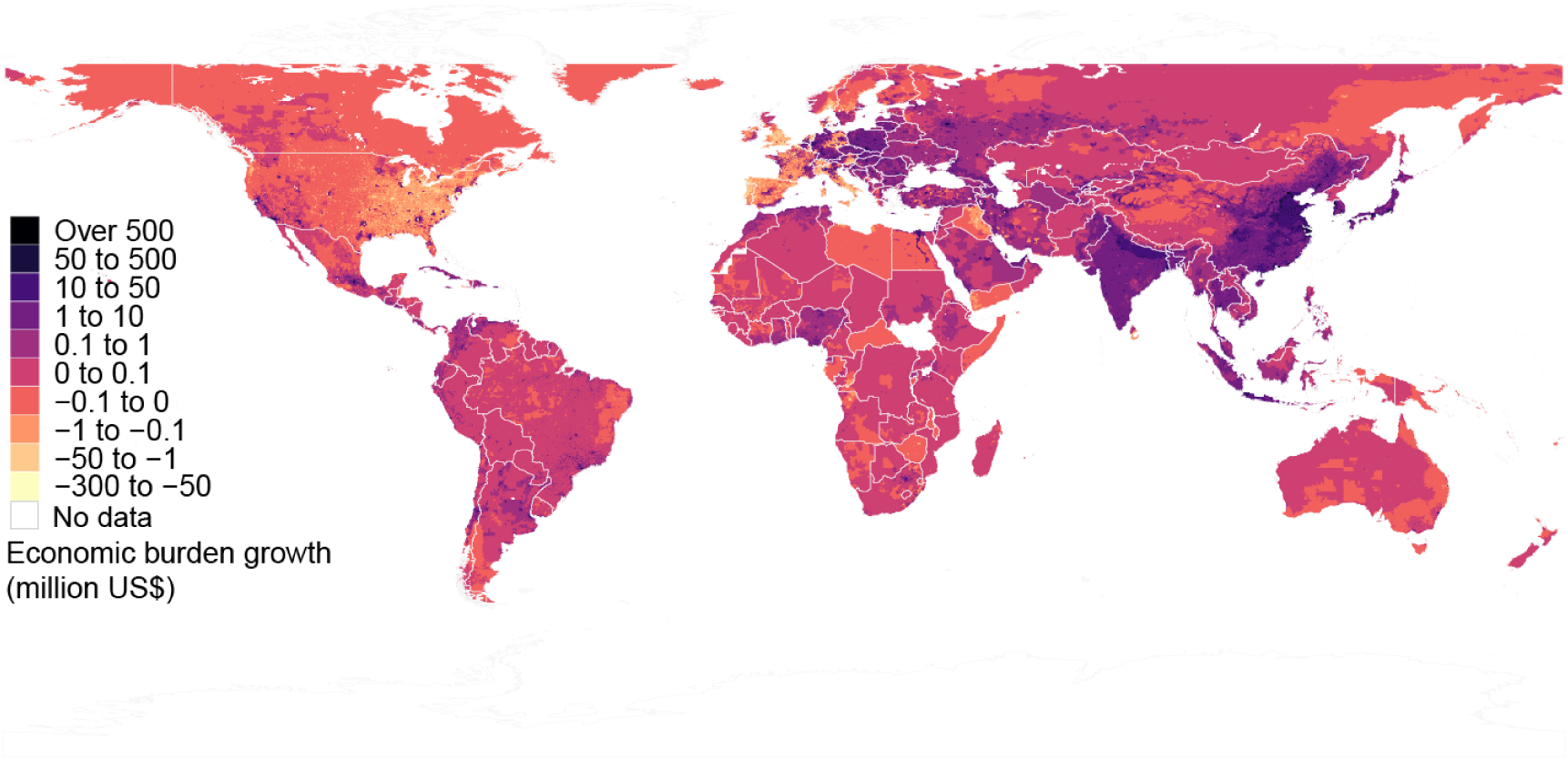
Growing health economic burden for the aged population from 2000 to 2016.

### 8. Health economic cost distribution with PM_2.5_ concentration

Figure 5a indicates the dramatic difference of health economic burden distribution with PM_2.5_ concentration by selected countries and the rest of the world. For high-income countries, such as the United States, Japan and Russia, the health cost mainly incurred in regions with PM_2.5_ concentrations below 25 μg/m^3^. By contrast, China and India suffered major economic loss with pollution concentrations between 25 μg/m^3^ and 100 μg/m^3^.

**Figure 5a.**
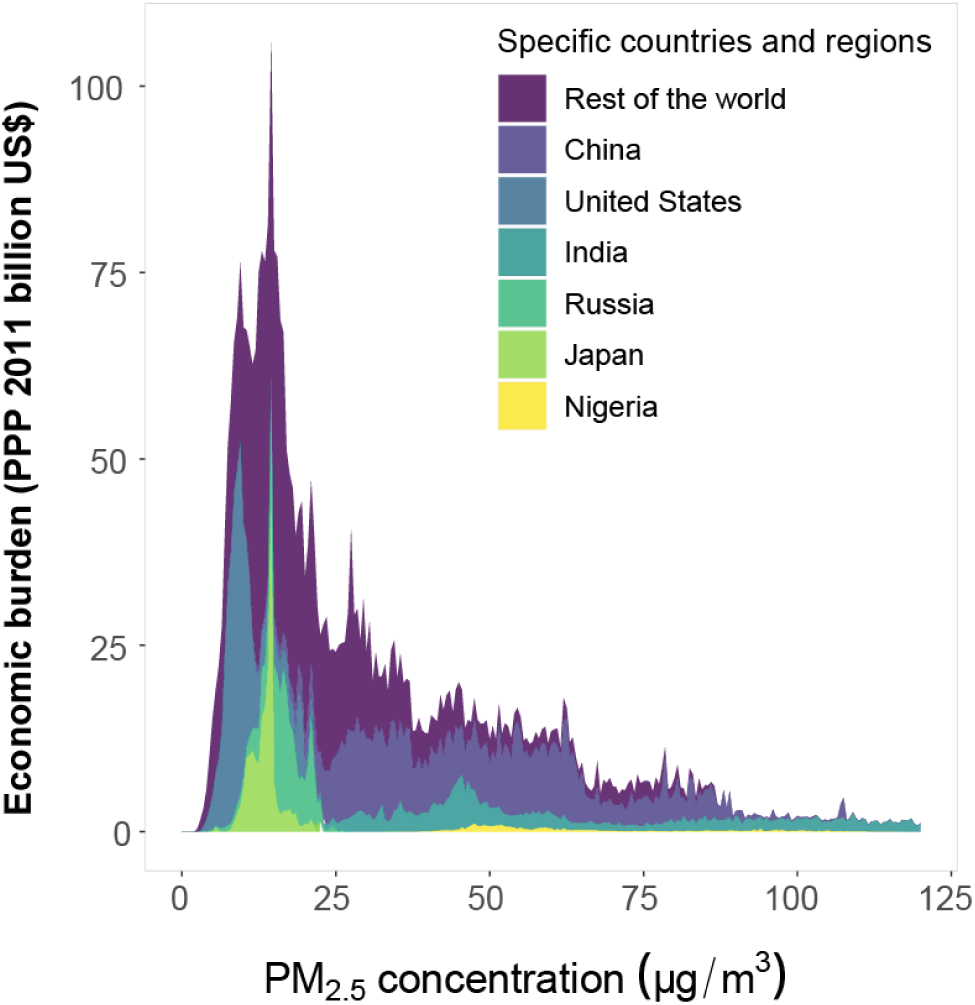
Economic burden distribution with respect to annual average PM_2.5_ concentrations in the six specific countries and the rest of the world.

### 9. Premature deaths, YLL and health cost by age and income group from 2000 to 2016

Figure 6a-8a demonstrate the variation of premature deaths, YLLs and health economic cost by age groups from 2000 to 2016. The lower-middle income countries suffered the highest premature deaths and YLLs, whereas the health economic cost related to such health impacts were lower than the loss in high-income countries. This is mainly because in lower-middle income countries costs for health risk abatement are lower.

**Figure 6a.**
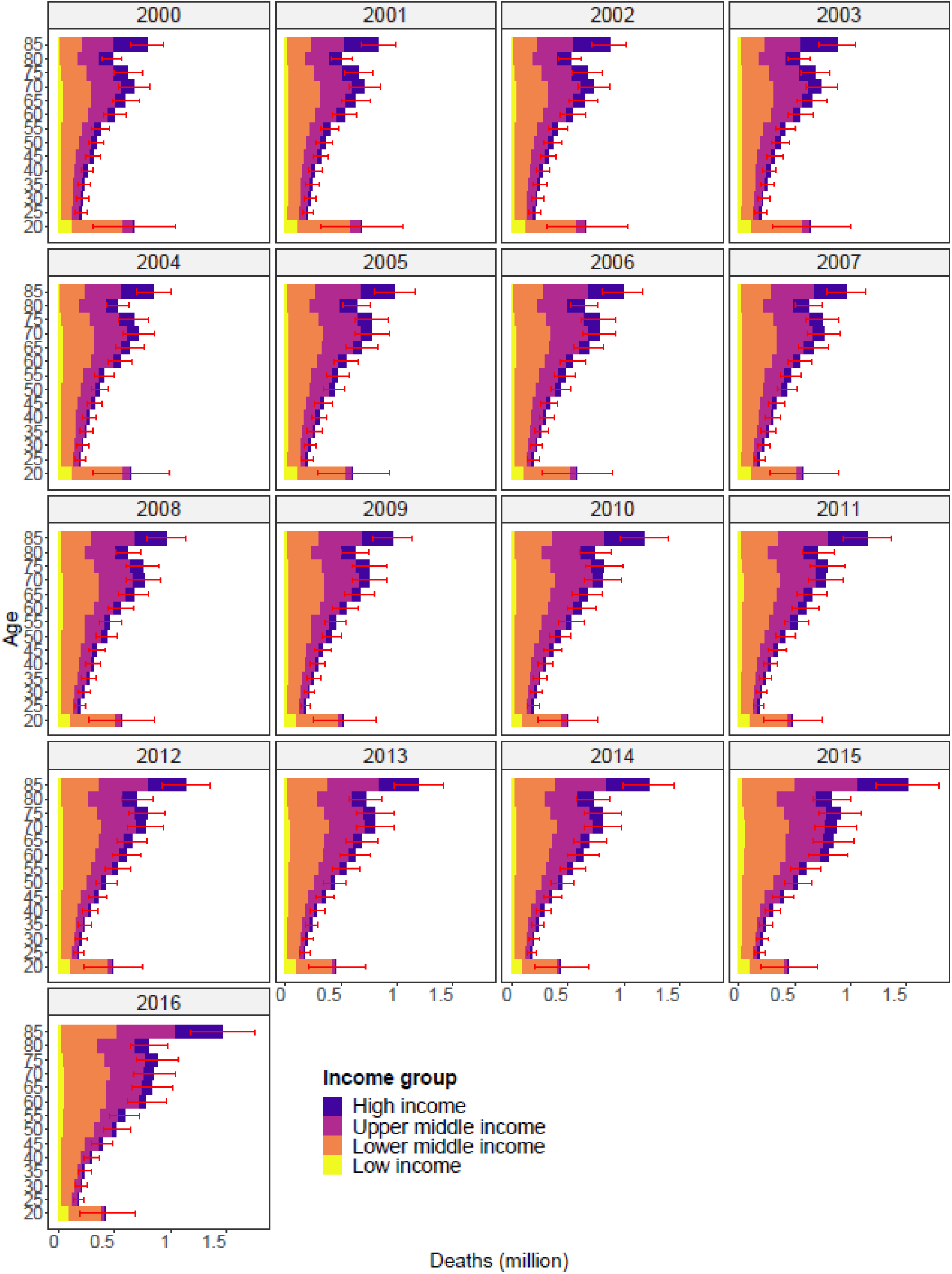
The attributable premature deaths from 2000 to 2016 by age in different income groups.

**Figure 7a.**
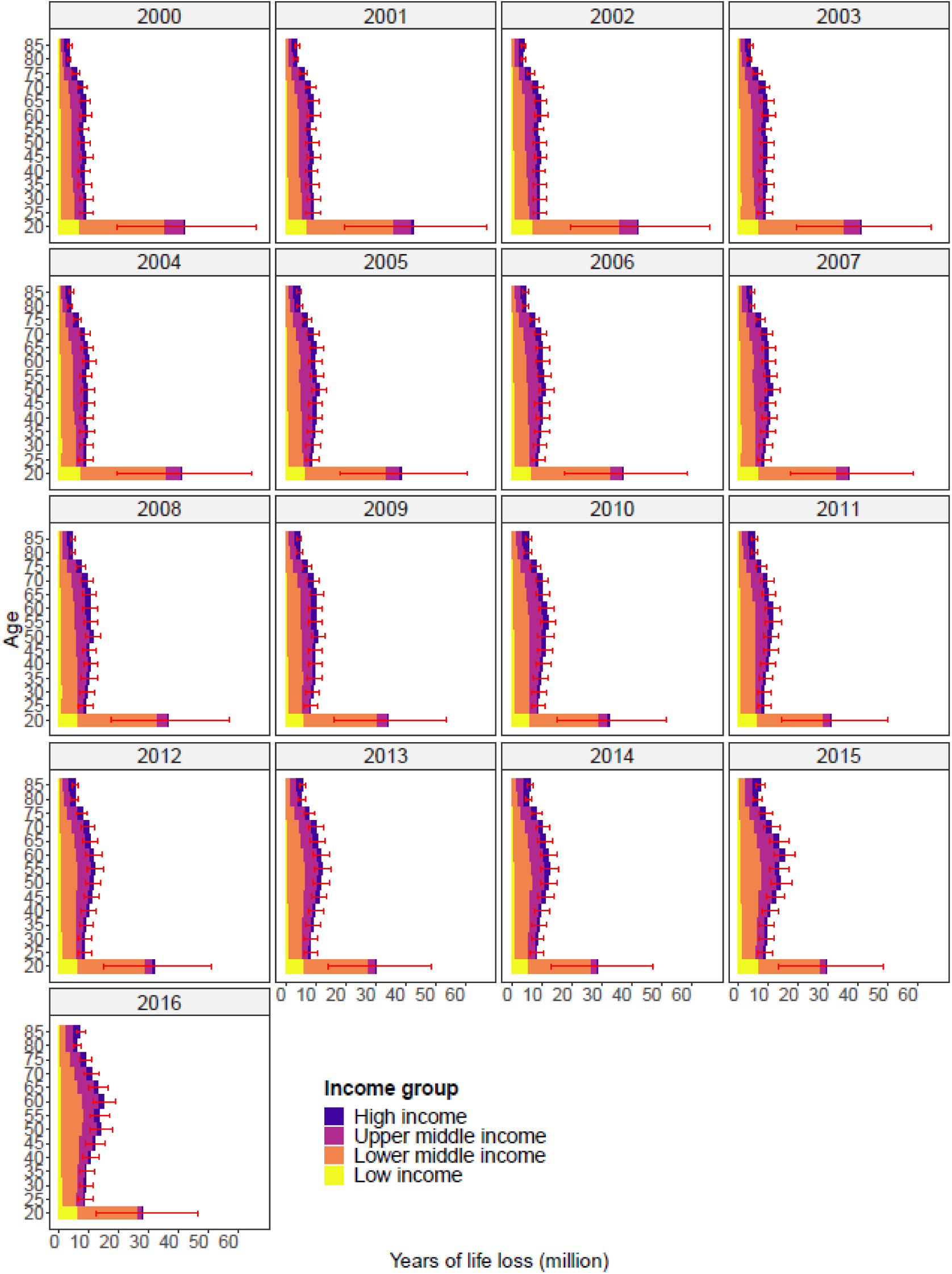
The attributable years of life lost from 2000 to 2016 by age in different income groups.

**Figure 8a.**
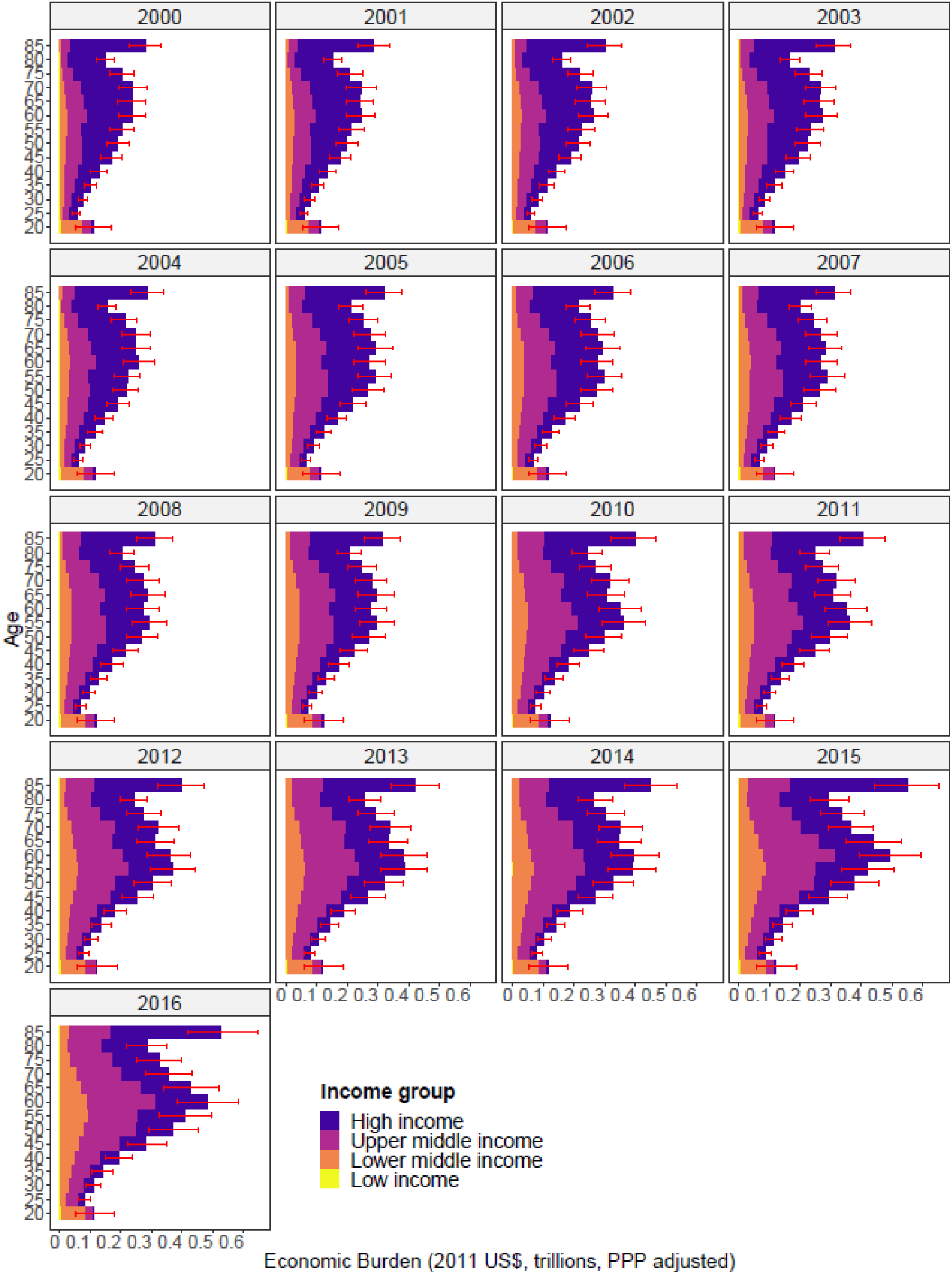
The attributable economic burden from 2000 to 2016 by age in different income groups.

### 10. Health economic cost trends by cause and GBD super regions

The health economic burden by causes and by regions are compared in Figure 9a. The Southeast Asia, East Asia, and Oceania region suffered 49% of global costs caused by COPD, followed by South Asia where it accounted for 21%. For lung cancer, Southeast Asia, East Asia, and Oceania and high-income regions experienced 47% and 36% of global health costs. High-income regions accounted for the highest proportion of total health costs caused by NCD-LRI and LRI. Southeast Asia, East Asia, and Oceania suffered the largest proportion of global health economic cost due to lung cancer, stroke and COPD.

**Figure 9a.**
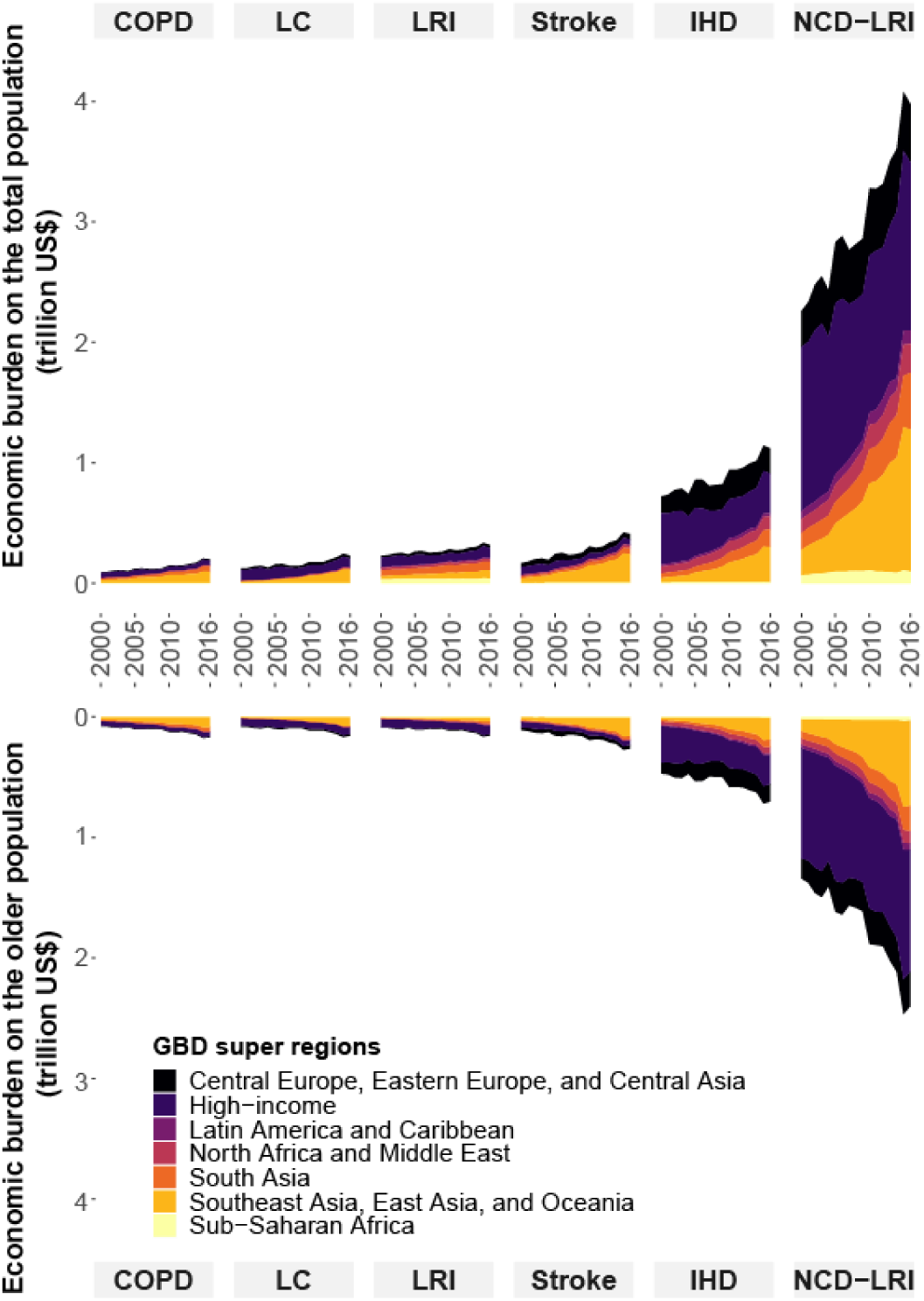
Comparison of health economic burden by different causes on the total and the older population between 2000 and 2016.

### 11. Cumulative health economic cost by valuation measure

Figure 10a illustrates the cumulative health economic burden distribution with respect to PM_2.5_ concentrations using five valuation measures. The age-invariant measure generates the highest estimates of economic cost related to PM_2.5_ pollution. With a global average age-adjusted VSLY measure, the estimates of economic burden grew substantially compared with country- & age-adjusted VSLY measure, which indicates that the most health damage happened in lower income countries.

**Figure 10a.**
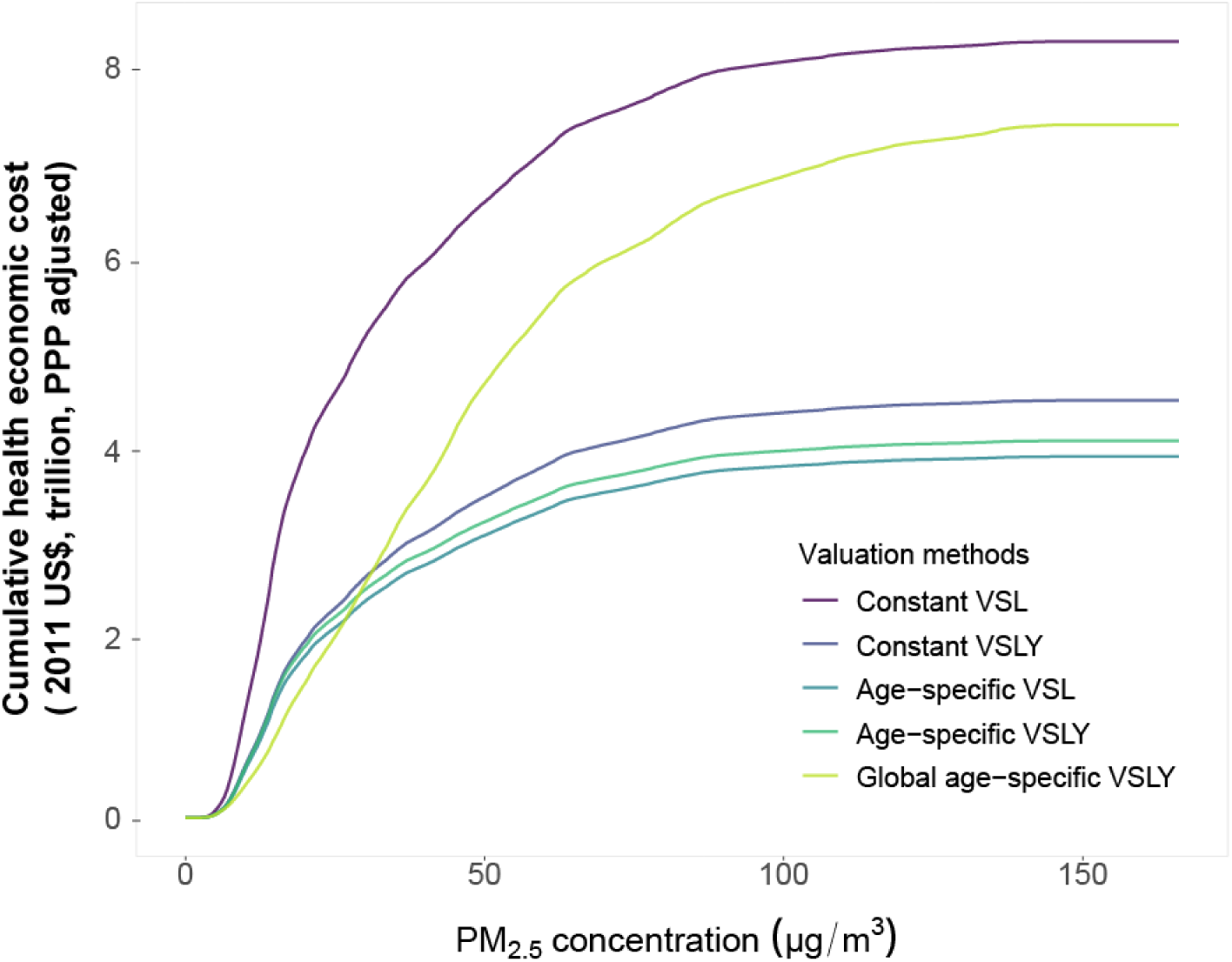
The growth cumulative health economic cost as a function of PM_2.5_ concentration by different valuation measures.

### 12. Cumulative health economic cost variations by age and measure

To further compare the differences in estimates, we disaggregated the health economic burden distribution by different age groups using five valuation measures. It shows that the economic burden difference among the population younger than 60 years old was relatively consistent. However, the gap became larger at 60 and older, and especially large above 85. Specifically, the health economic burden valued by age-invariant VSL was 4.3 times the loss estimated by age-adjusted VSLY on the population aged 85 and higher.

**Figure 11a.**
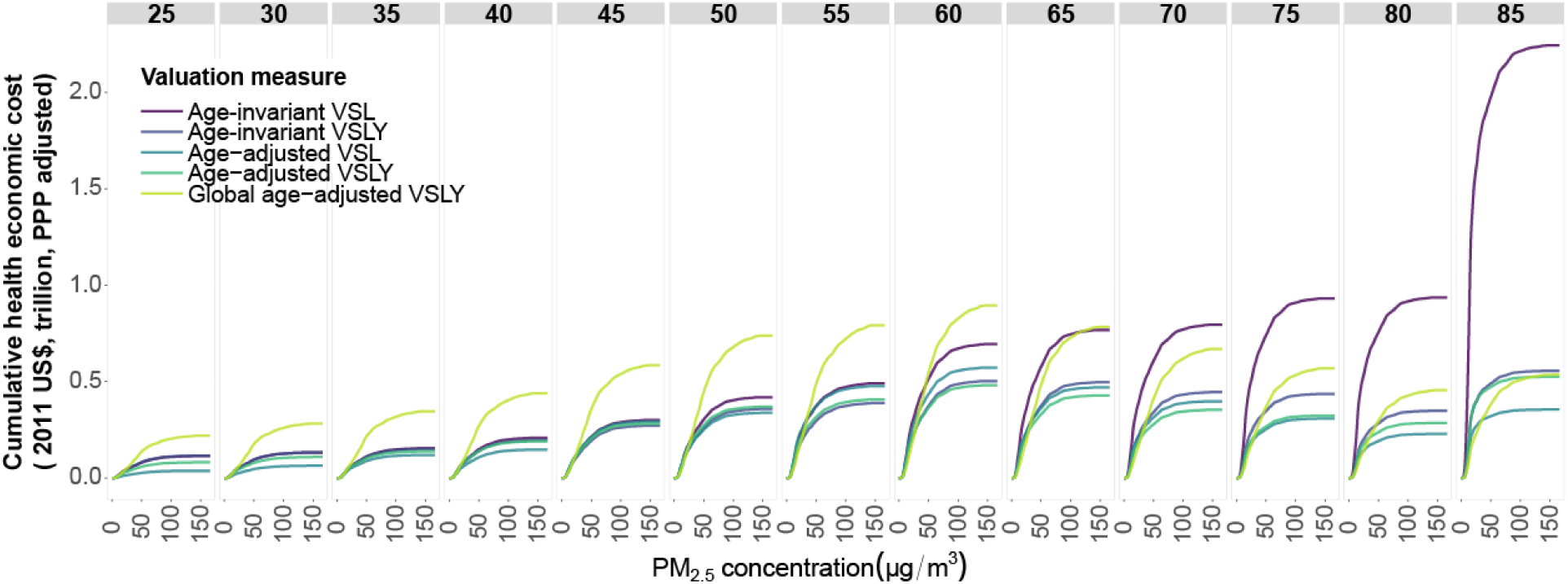
The comparison of cumulative economic burden distribution with PM_2.5_ concentration by age using five valuation measures.

### 13. Age-VSL pattern of different valuation measures

Figure 12a illustrates the effect of age on VSL over a life cycle period. In the age-adjusted VSL/VSLY measures, there were two inverted U-shape patterns after 20 years old, which shows that the VSL peaked at 40–50 years old (Figure 11a), which is consistent with previous studies ^4^ Since people younger than 20 years old generally have little cumulative wealth and there is no such data available for the analysis, we assumed the wealth weights are similar to their parents’ wealth weights. For example, the wealth weight for people at 0–5 years old matched with wealth data of their parents aged around 25–30 years old. Therefore, the inverted U-shape pattern before 20 years old is due to the inverted relationship between age and VSL in the population at working age. Many empirical wage-risk studies revealed an inverted U-shaped pattern of age-VSL relationship ^5,6^. However, the hedonic wage models only revealed the WTP for mortality risk reduction during one’s working age, i.e., in a rough range of 18 to 62 years old ^4^ Due to limited studies reported the age-VSL relationship for people under 20 years old, we assumed that people under 20 years old have the same wealth as that of their parents.

**Figure 12a.**
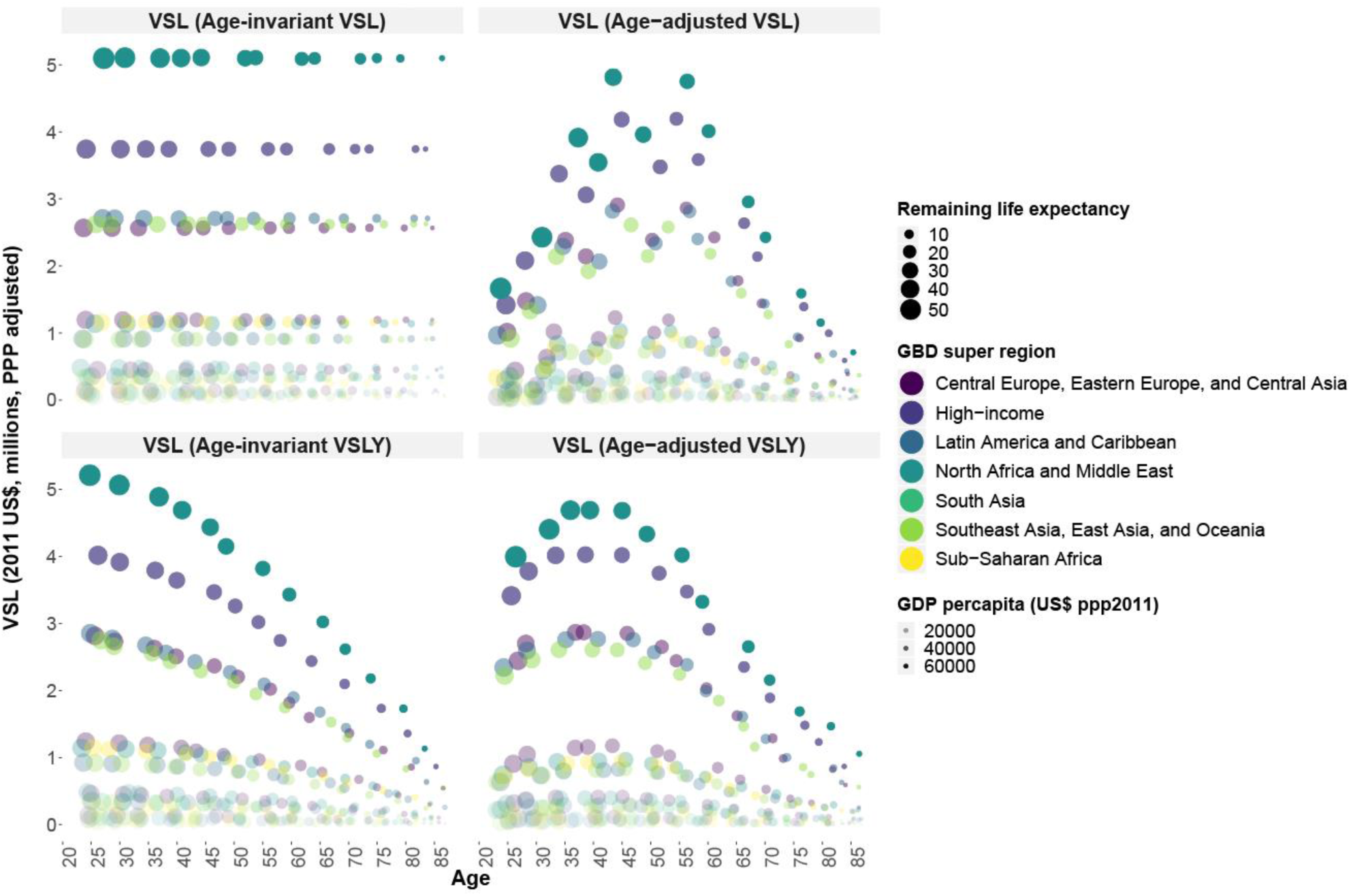
Age-VSL patterns over a lifetime.

